# Role of Chromosomal Microarray and RNA Fusion Analysis in Detecting *KMT2A*-PTD and Stem Cell Transplant Impact on Mortality

**DOI:** 10.1101/2025.05.27.25328455

**Authors:** Bhaumik Shah, Roniya Francis, Ashishkumar Sonani, Jianming Pei, Peter Abdelmessieh, Mariusz A. Wasik, Nicholas Mackrides, Reza Nejati

**Author notes:** Corresponding author: Reza Nejati, Department of Pathology, Fox Chase Cancer Center, Temple University Health System, 333 Cottman Avenue, Philadelphia, PA 19111;.

## Abstract

*KMT2A* partial tandem duplication (*KMT2A*-PTD) is a recurrent, high-risk alteration in myeloid neoplasms, yet no gold standard exists for its detection due to complex genomic architecture. We conducted a retrospective study of 97 specimens from 17 patients with *KMT2A*-PTD-positive myeloid neoplasms (11 AML, 4 MDS, 2 MPN) to compare chromosomal microarray analysis (CMA) and RNA fusion testing. Overall concordance was 73.3% (κ = 0.467), with RNA fusion identifying more cryptic *KMT2A*-PTDs (p = 0.035), while CMA detected non-canonical *KMT2A*-PTDs and, additionally, secondary genomic abnormalities (e.g., trisomy 8 and CN-LOH of diverse genes). *KMT2A-*PTD cases exhibited high clonal complexity and distinct mutational profiles, including mutations of *DNMT3A*, *TET2, ASXL1, RUNX1, FLT3*, and others. Higher *KMT2A*-PTD transcript expression (split reads and SR/C ratio) correlated with *DNMT3A*, *RUNX1*, and *FLT3* mutations, and *RUNX1T1* copy number gain (p < 0.05), and was reduced in CN-LOH cases. RNA fusion consistently identified breakpoints at exon 10→2 or exon 11→2, affecting key oncogenic domains (CXXC, PHD finger, and bromodomain). Six-month overall survival was 41.2%, with HSCT significantly improving outcomes (68.6% vs. 0% at 17 months, p = 0.028), non-transplanted patients showing shorter median PFS (∼6 months), and *FLT3*-mutated patients experiencing 100% mortality (p = 0.0456). In conclusion, CMA and RNA fusion analyses offer complementary diagnostic value in *KMT2A-PTD-positive myeloid neoplasms.* Such integrated genomic testing improves detection and refines risk stratification. Given the poor prognosis associated with *KMT2A*-PTD, especially in patients with co-occurring high-risk mutations, our findings support early consideration of HSCT in such patients.

## Introduction

Partial tandem duplication of *KMT2A* gene (*KMT2A*-PTD) is a recurrent genetic alteration in myeloid neoplasms, seen in 5-10% of acute myeloid leukemia (AML), 2–7% of myelodysplastic syndromes (MDS), and, rarely, in myeloproliferative neoplasms (MPN) [1, 2]. This structural variant, typically spanning 10-50 kb, involves in-frame duplication of *KMT2A* exons (e.g., 2-8 or 2-10) via Alu-element-mediated homologous recombination, disrupting the CXXC and PHD domains [1]. The resultant aberrant protein drives leukemogenesis by upregulating *HOX* genes, promoting cell proliferation and differentiation arrest, linked to poor prognosis [3]. Approximately 16% of cases develop secondary genomic events, such as copy-neutral loss of heterozygosity (CN-LOH) or *KMT2A*-PTD allele amplification, enhancing RNA expression and promoting disease progression. *KMT2A*-PTD frequently co-occurs with mutations in *DNMT3A*, *RUNX1*, and *FLT3* genes, amplifying epigenetic and cell-signalling dysregulation, raising the risk of leukemic transformation in MDS/MPN, and increasing relapse rates [4, 5]. Its expression and complexity are notable in high-risk/advanced MDS or MPN, or secondary AML. Clinically, *KMT2A*-PTD confers a poor prognosis, (median overall survival (OS) 9.85 months in MDS, 4.5–12.1 months in AML), driven by chemotherapy-resistance and relapse [4, 6]. It also serves as a biomarker for menin inhibitors, targeting the menin-*KMT2A* interaction, offering a novel therapeutic avenue. In contrast to *KMT2A* rearrangements, which are well characterized in acute leukemias and have established diagnostic and therapeutic guidelines, *KMT2A*-PTD remains under-recognized, with no consensus gold standard for detection and limited data on its clinical significance and management. This unmet need reinforces the importance of developing and validating optimal diagnostic approaches to optimize risk stratification and therapeutic decisions for this high-risk group. Recent advances in understanding *KMT2A* allelic complexity [8] and consensus gene structure [9] have highlighted the need for precise genomic mapping, while epigenetic alterations induced by *KMT2A* partial tandem duplications underscore their leukemogenic potential [13].

*KMT2A*-PTDs are complex gene rearrangements that cannot be fully ascertained using a single genomic platform. Its detection is challenging due to small size, below the resolution limits of karyotyping (5–10 Mb) or standard FISH *KMT2A* break-apart probes [7]. Furthermore, its intragenic nature leads to significant overlap in interphase nuclei making its detection unreliable, even with custom probe sets targeting the *KMT2A*-PTD region. Optical genome mapping (OGM) can resolve large structural variants, such as 11q gains associated with *KMT2A*-PTD, but is less sensitive for small duplications [7]. Several methods, including chromosomal microarray (CMA), RNA fusion analysis (e.g., Illumina TruSight RNA Fusion Panel), multiplex ligation-dependent probe amplification (MLPA), next-generation sequencing (NGS), and reverse transcription PCR (RT-PCR), are used, each with distinct limitations **(Supplemental Table 1).** Long-range RT-qPCR, was historically considered the gold standard and targeting canonical PTDs (exons 2–8 or 2–10), is limited by exon specific primer dependence, inability to detect non-canonical exons or cryptic events and the influence of complex genomic structures or fusion variants during disease progression—limits its value as a true gold standard in clinical settings [2]. Routine NGS panels, optimized for short nucleotide variants, require custom pipelines for reliable detection due to poor intronic coverage and high VAF thresholds [5, 11]. Hybrid capture NGS assays (e.g., Oncomine) can detect *KMT2A*-PTDs and co-mutations at variant allele frequencies of at least 10%, but also requires customized complex bioinformatics pipelines, limiting its potential as a gold standard in routine clinical detection [2].

Chromosomal microarray (CMA) offers an alternative by detecting copy-number alterations (CNAs) and copy-neutral loss of heterozygosity (CN-LOH), but is limited for small intragenic duplications without CNAs [7]. RNA fusion panels, using split-read analysis, detect fusion transcripts with high sensitivity, even in cases with low RNA quality, making them particularly valuable for residual disease monitoring [5]. No study has directly compared CMA and RNA fusion panels for *KMT2A*-PTD detection, despite their complementary strengths. In this retrospective study of 97 specimens across 17 patients with *KMT2A*-PTD myeloid neoplasms at Fox Chase Cancer Center (2011 – 2025), we provide the first head-to-head evaluation of CMA and RNA fusion panels, comprehensively assessing their diagnostic performance. Additionally, we analyzed survival outcomes, co-mutations via NGS, and additional abnormalities (FISH, karyotype, *FLT3*) to elucidate prognostic and therapeutic implications of *KMT2A*-PTD, highlighting the impact of hematopoietic stem cell transplantation (HSCT) in reducing mortality and informing optimal detection strategies and patient management for this high-risk cohort.

## Methods

### Study Design and Specimen/Patient Selection

This retrospective study reviewed molecular and cytogenetic data from the Fox Chase Cancer Center database to identify specimens with confirmed *KMT2A*-PTD) between January 2011 and April 2025. Search criteria included specimens with *KMT2A*-PTD detected by chromosomal microarray analysis (CMA) and/or RNA fusion panels, along with a concurrent diagnosis of myeloid neoplasm (AML, MDS, or MPN) at the time of *KMT2A*-PTD detection.

Inclusion criteria required *KMT2A*-PTD to be detected by CMA and/or RNA fusion panel; exclusion criteria were age under 18 years, non-myeloid neoplasms, or unconfirmed *KMT2A*-PTD. Our cytogenetic database comprised 5,218 eligible specimens analyzed with CMA (Affymetrix CytoScan HD, 2,696,168 markers) and 720 eligible specimens analyzed with NGS-based targeted RNA sequencing (Illumina TruSight RNA Fusion 523-gene panel). Corresponding bone marrow and peripheral blood specimens were verified in the pathology database to determine the total number of unique patients with confirmed *KMT2A*-PTD. Patients were grouped based on their diagnosis at the time of *KMT2A*-PTD detection (AML, MDS, or MPN). To enable direct comparison between CMA and RNA fusion methods in absence of definitive gold standard test, all available longitudinal specimens were included, for cross-validation. The study was approved by the Institutional Review Board and conducted in accordance with the Declaration of Helsinki.

### Data Collection

Patient demographics, clinical history (diagnosis/ disease duration, mortality, transplant status, cause of death) and survival data wereextracted from the pathology database and EMRs. Overall survival (OS) was defined as the time from *KMT2A*-PTD detection (typically at disease progression) to death from any cause or to the last follow-up (April 30, 2025). PFS was defined as the time from the relevant baseline (*KMT2A*-PTD detection for non-transplant patients, or transplant for transplant recipients) to the first occurrence of death from any cause or relapse (≥5% blasts following achievement of complete remission). Patients without an event were censored at last follow-up. Ancillary data, including NGS-based targeted DNA sequencing (275-gene Comprehensive Cancer Panel v1), FISH, karyotyping, *FLT3*, and *NPM1* studies, were extracted to resolve discordant CMA and RNA fusion results using a composite reference standard. NGS, was previously performed previously as part of clinical work up, utilized the 275-gene Comprehensive Cancer Panel v1 (Illumina), including myeloid-relevant genes (e.g., *DNMT3A, FLT3, RUNX1*) as part of the clinical workup.

### Genomic Coordinate Verification and *KMT2A*-PTD Classification

Genomic coordinates (from CMA/RNA reports) were verified using the UCSC Genome Browser (hg19 assembly) referencing the ENST00000534358.1 transcript (GRCh37.p13, *KMT2A*-001; Chromosome 11:118,436,492–118,526,832, forward strand) [9]. This process determined the precise genomic coordinates, duplicated exons and introns, PTD size, and affected functional domains (e.g., CXXC, PHD, and SET). This approach was informed by detailed cytogenetic studies of *KMT2A* rearrangements [14], ensuring accurate delineation of structural variants. *KMT2A*-PTD cases were classified as simple (net +1 copy gain of PTD exons) or complex (e.g., +2/+3 copy gains, CN-LOH, trisomy 11) based on allelic state, copy number, and VAF, adopting definitions from Tsai et al. (2022) [8](Table 1). Quantitative *KMT2A*-PTD expression was measured using total split reads and split reads per coverage (SR/C) ratio (Supplemental Table 5) and categorized as high, moderate or low.

**Table 1.** Definition of Simple and Complex *KMT2A*-PTD Allelic States and Copy Number Variations (adopted from Tsai et al. (2022)).

| Type | Allelic State | Copy Number | Example |
| --- | --- | --- | --- |
| Simple<br>*KMT2A*-PTD | 1 wild-type allele (1 copy of *KMT2A*) + 1 PTD allele (2 copies of PTD exons, 1 copy of non-PTD exons) | Net +1 copy gain of PTD exons (total 3 copies), 2 copies of non-PTD exons | Diploid cell with one PTD allele, typically mosaic (VAF <100%), no trisomy 11 or CN-LOH on 11q23.3 |
| Complex<br>*KMT2A*-PTD |  |  |  |
| - Gain of 11q23.3 | 1 wild-type + 2 PTD alleles | +3 copy gain of PTD exons (5 copies), +1 copy gain of non-PTD exons (3 copies) | Trisomy 11 or multiple PTD alleles |
| - Double Gain of 11q23.3 | 1 wild-type + 3 PTD alleles | +5 copy gain of PTD exons (7 copies), +2 copy gain of non-PTD exons (4 copies) | Multiple PTD alleles or additional 11q23.3 gains |
| - CN-LOH | 0 wild-type + 2 PTD alleles | +2 copy gain of PTD exons (4 copies), no gain of non-PTD exons (2 copies) | CN-LOH on 11q23.3 |
| Log2 Ratio Calculation | For VAF ( *f* ), copy number = ( 2 + *f* ), Log2 Ratio = ( log<sub>2</sub>((2 + *f*)/2) | Confirms copy number gain of PTD exons relative to diploid baseline | Used to quantify copy number changes in genomic analysis |

Note: The Log2 Ratio calculation quantifies copy number gain of PTD exons based on VAF, enabling comparison to a diploid baseline for genomic analysis.

### Assessment of Diagnostic Performance of CMA and RNA fusion analysis

A composite reference standard (CRS) was constructed to classify specimens as *KMT2A*-PTD-positive or *KMT2A*-PTD-negative, integrating CMA, RNA fusion analysis, clinical correlation, bone marrow morphology, and ancillary findings (NGS, FISH, karyotyping). The specimen was positive if CMA detected 11q23.3 gains, or RNA fusion analysis identified a fusion transcript. True positives (TP), false negatives (FN), true negatives (TN), and false positives (FP) were adjudicated pre– and post-CRS (Supplemental Methods).

## Statistical Analysis

The diagnostic performance (sensitivity, specificity, PPV, NPV) was assessed pre– and post-CRS. Concordance was quantified using overall concordance, *KMT2A*-PTD-specific concordance, Cohen’s Kappa, and McNemar’s test. Co-mutation frequencies were determined from NGS data, with associations tested using Fisher’s exact test. *KMT2A*-PTD expression was correlated with mutations and outcomes using Mann-Whitney U and Kruskal-Wallis tests. PFS and OS were analyzed using Kaplan-Meier curves and log-rank tests, stratified by diagnosis, transplant status, and mutation status.

## Results

### Patient demographics and Clinical Characteristics

The study identified 17 patients (12 males, 5 females; median age 68 years, range 42–79) with *KMT2A*-PTD-positive myeloid neoplasms, comprising 11 AML (6 secondary [transformed from MDS or MPN], 5 *de novo*), 4 MDS (2 high grade), and 2 MPN (1 high grade) **(Table 2)**. HSCT was performed in 7 patients (41.18%). Median time from *KMT2A*-PTD detection to transplant was 5 months, with a median disease-free survival post-transplant of 12 months.

**Table 2.**
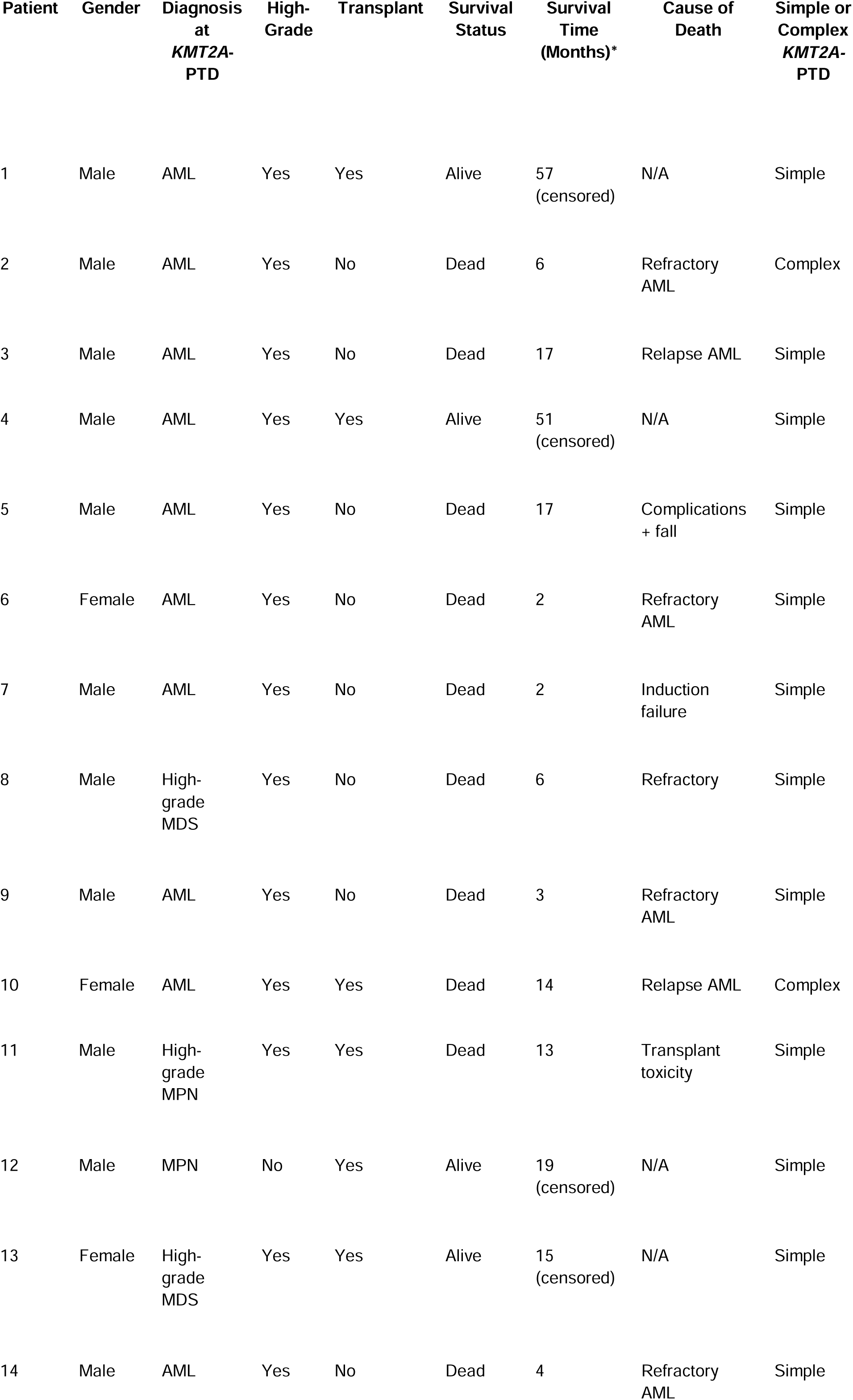
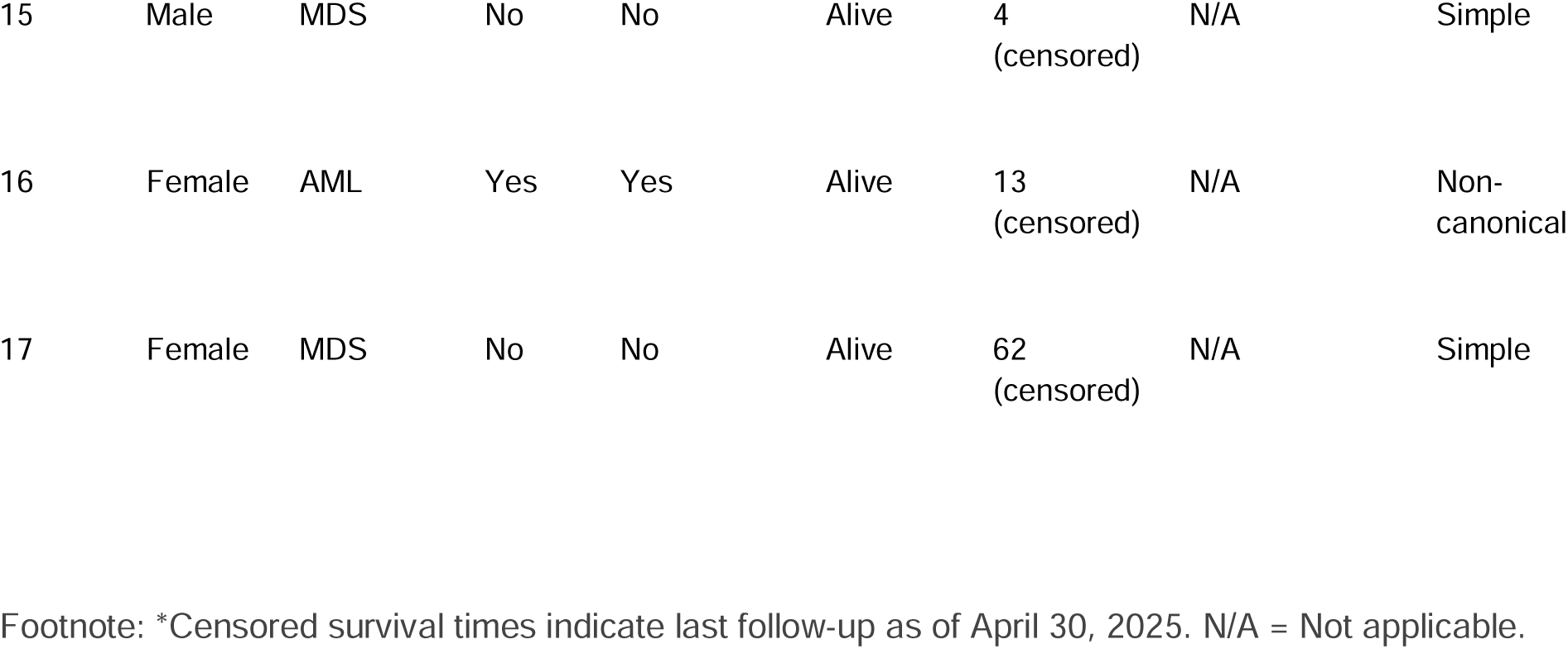
Patients’ clinical characteristics, mortality status and *KMT2A*-PTD status.

### Specimen characteristics

Of 97 specimens (79 bone marrow, 18 peripheral blood), 31 were *KMT2A*-PTD positive, detected by CMA (24/31) and/or RNA fusion (21/22). *KMT2A*-PTD was classified as simple (14 patients: 2, 3, 4, 5, 6, 7, 8, 9, 11, 12, 13, 14, 15, 17), complex (2 patients: patient 2 [trisomy 11, +2 *KMT2A*-PTD copy gain] and patient 10 [11q CN-LOH, +2 the copy gain]), or non-canonical (1 patient: 16, exons 27–36) **(Table 1).** The CRS identified 35 PTD-positive specimens’ post-adjudication, including 4 additional false negatives from concordant/discordant negatives identified by NGS, bone marrow morphology and/or FISH. These included three discordant specimens that were negative by CMA without RNA fusion analysis, as well as one discordant specimen that was negative for RNA fusion, with CMA not performed. The specimens negative for *KMT2A*-PTD by CMA (n=3) showed bone marrow morphology demonstrating 8% blasts (e.g., Patient 4), 10–14% blasts (Patient 2), or 6% circulating blasts (Patient 11). In the specimen that was negative for RNA fusion (Patient 16), FISH demonstrated 8% blasts.

### Diagnostic Performance

Pre-CRS, CMA detected *KMT2A*-PTD in 24/31 specimens (sensitivity 77.42%, specificity 100%, PPV 100%, NPV 76.67%), while RNA fusion detected 21/22 (sensitivity 95.45%, specificity 100%, PPV 100%, NPV 90.91%). Post-CRS, CMA sensitivity decreased to 68.57% (24/35) and RNA fusion to 87.50% (21/24) due to additional false negatives (e.g., Patient 4’s 8% blasts, Patient 2’s 10–14% blasts), with specificity and PPV remaining 100% (NPV: CMA 63.33%, RNA 72.73%). Both CMA and RNA fusion analysis demonstrated 100% specificity and positive predictive value (PPV), as no false positives were identified due to validation by the composite reference standard, ensuring all positive results were true positives.

Overall concordance was 73.33% (κ=0.4667), with 45.16% *KMT2A-PTD* specific concordance. RNA fusion outperformed CMA in discordant cases (p=0.03516). *KMT2A* gain was detected by FISH in 5 of 22 specimens (16.13% sensitivity) and by karyotype in 7 of 37 specimens (22.58% sensitivity). *FLT3* mutations were found in 9 of 19 specimens from 4 patients (100% sensitivity).

### Chromosomal Microarray Analysis

CMA confirms *KMT2A*-PTD in Patients 1, 2, 3, 8, 9, 10, 11, 12, 13, 14 (partial), 15, 17, with sizes (10,890–65,301 bp) overlapping RNA breakpoints. Patients 4–7 lacked detection by CMA, but were detected by RNA fusion panel, indicating cryptic PTDs, despite high blast percentage in the specimens. *KMT2A*-PTD genomic coordinates reported by CMA (**Figure 1**) were mapped to the ENST00000534358.1 transcript (*KMT2A*-001; GRCh37.p13, hg19), confirming duplicated exons (e.g., 2–8, 2–10) (Figure 2) and functional domains (CXXC, PHD, and SET) (**Supplemental Table 2**). Patient 16 showed an atypical *KMT2A*-PTD involving introns 26-27 to 35-36 and exons 27-36 (SET domain), outside the typical breakpoints in exons 2-11, raising questions about its classification as a true *KMT2A*-PTD High-frequency region (Exons 2–5, Introns 1-2 to 4-5) was covered in 17 of 20 specimens, indicating a core region of *KMT2A*-PTDs affecting the CXXC domain (exons 4–5), critical for oncogenic activity in AML, MDS, and MPN. Mid-frequency region (exons 6–8 and introns 6-7 to 8-9) involved the PHD domain (exons 6–10) covered in 8 specimens (patient 1, 2, 9, 11, 13, 14, 15, and 17), and the AT Hooks (exons 1–2) in five cases (patient 2, 3, 11, 15, and 17). Involvement of the bromodomain (exons 11–20) was observed in three cases (patient 2, 14, and 17). The degree of CXXC domain involvement varied, with partial involvement noted in five cases (patient 1, 3, 8, 10, and 13).

**Figure 1.**
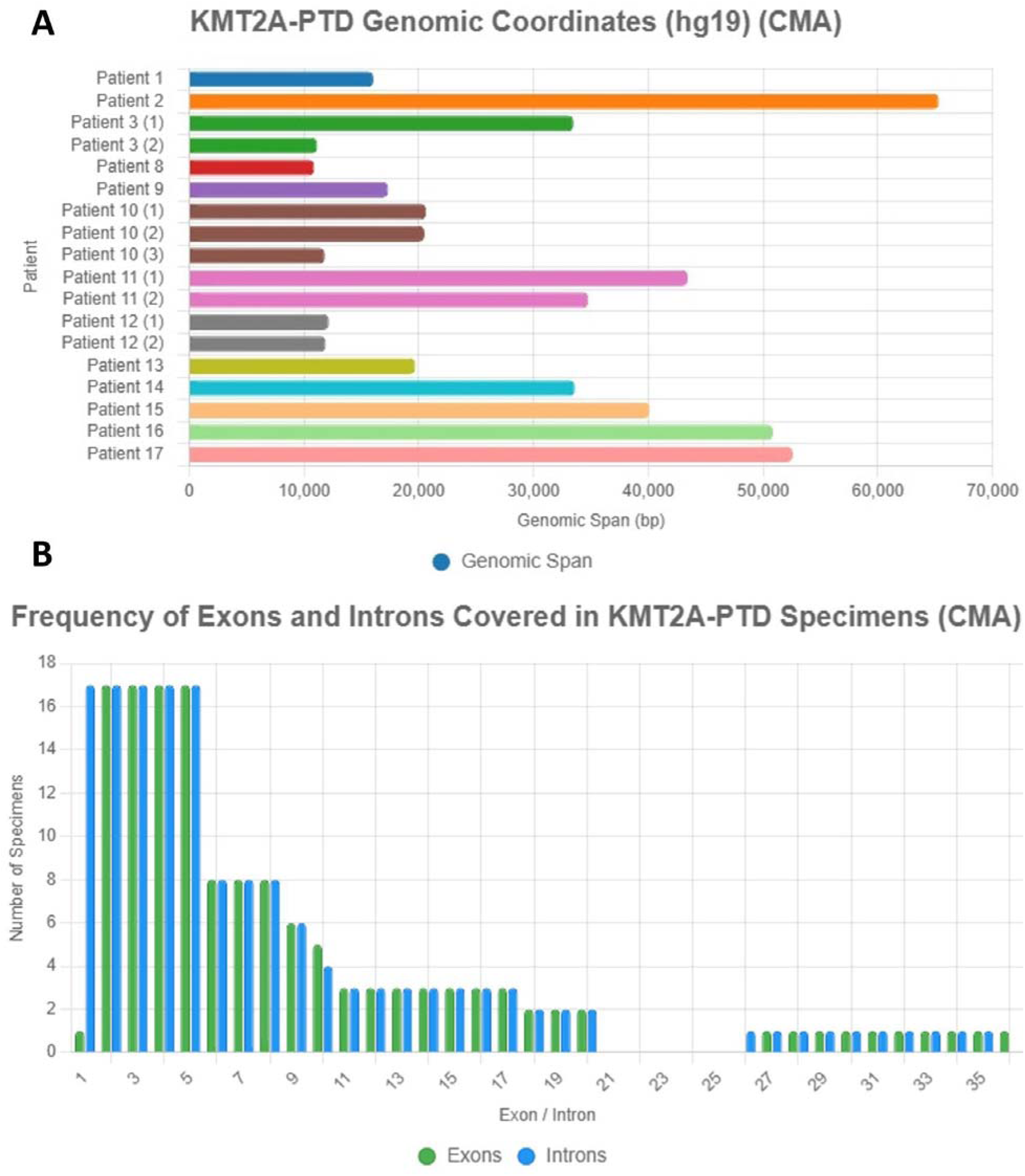
Genomic Coordinates and Exon/Intron Coverage of *KMT2A*-PTD Detected by Chromosomal Microarray (CMA). **(A)** Horizontal bar plot displaying genomic coordinates and sizes of *KMT2A*-PTD regions for 12 patients (1, 2, 3, 8, 9, 10, 11, 12, 13, 14, 15, 17) mapped to the ENST00000534358.1 transcript (GRCh37.p13, hg19). Bars range from 10,890 bp (Patient 8: 118,339,219–118,350,109) to 65,301 bp (Patient 2: 118,302,712–118,368,013), with Patient 16’s non-canonical PTD (50,853 bp, 118,374,566–118,425,419, exons 27–36, introns 26–27 to 35–36) in green. **(B)** Stacked bar plot showing exon (green) and intron (blue) coverage frequency across specimens, with high involvement in exons 2–5 (CXXC domain, ∼12 patients) and reduced in exons 6–10 (PHD domain, ∼8 patients). Patients 4–7, undetected by CMA but confirmed by RNA fusion, are excluded. The figure aids in identifying core PTD regions (exons 2–5) and functional domains (CXXC, PHD, SET) critical for leukemogenesis. Data reflects analysis up to April 30, 2025.

**Figure 2.**
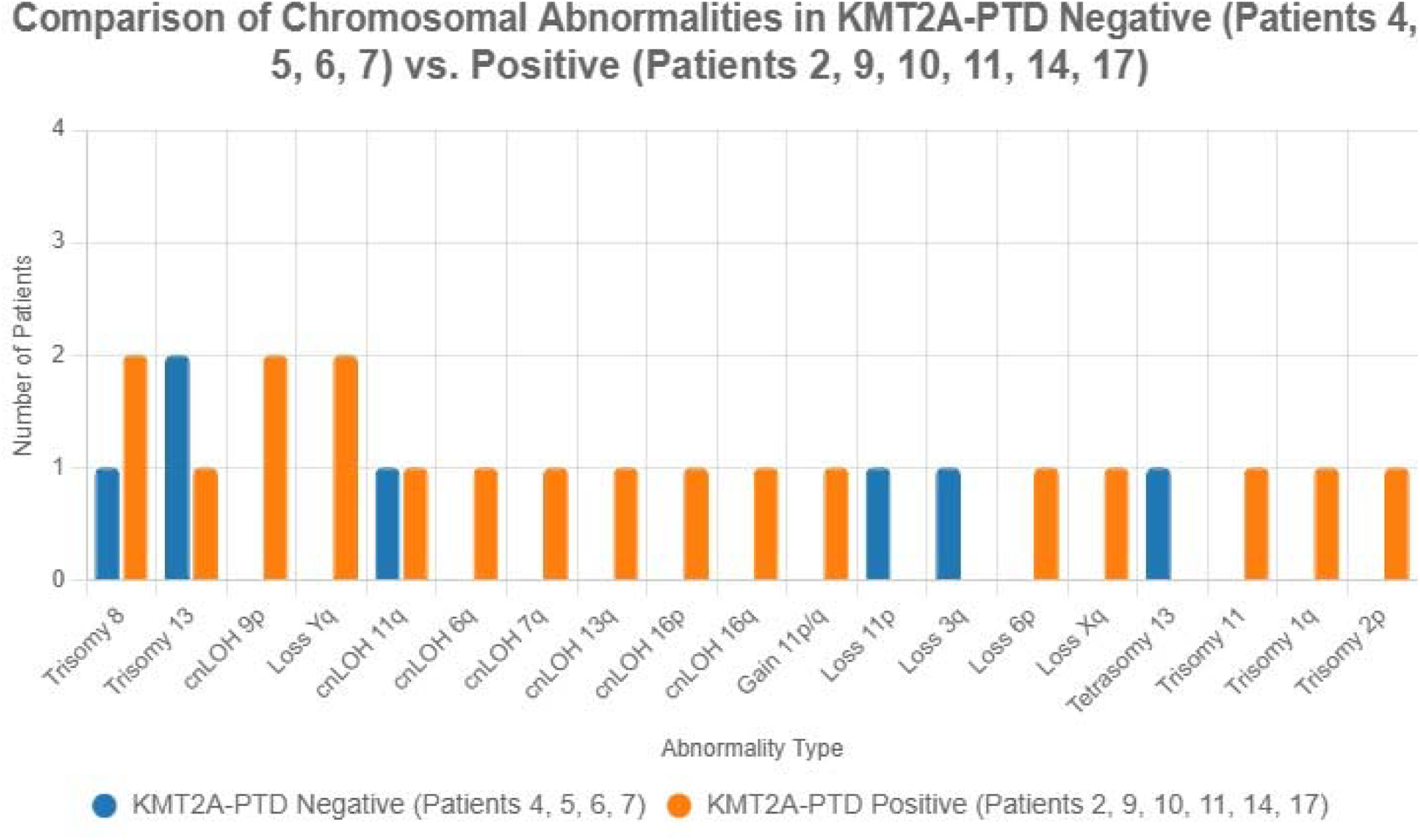
Comparison of Chromosomal Abnormalities in *KMT2A*-PTD-Negative versus *KMT2A*-PTD-Positive Groups. Bar chart comparing the frequency and types of chromosomal abnormalities detected by Chromosomal Microarray (CMA) in *KMT2A*-PTD-negative (Patients 4, 5, 6, 7; n=4) and *KMT2A*-PTD-positive (Patients 2, 9, 10, 11, 14, 17; n=6) groups, based on 97 specimens. The positive group exhibits 17 abnormality types (e.g., *KMT2A* gain [11q23.3, 3 copies], trisomy 11 [Patient 2], CN-LOH 7q [EZH2, Patient 14]) versus 6 in the negative group (e.g., loss 3q [Patient 6], trisomy 8 [Patients 5, 14]). Colors distinguish abnormality categories (e.g., gains, losses, CN-LOH). Higher complexity in the positive group correlates with increased mortality (83.3% [5/6] vs. 75% [3/4], data up to April 30, 2025). Coordinates are mapped to hg19 assembly.

#### Additional chromosomal abnormalities detected by CMA

In seven patients (1, 3, 8, 12, 13, 15, and 16), *KMT2A*-PTD was identified by CMA as the sole cytogenetic abnormality. Conversely, in four patients (4, 5, 6, and 7), while CMA did not detect *KMT2A*-PTD, other genomic aberrations were observed **(Figure 2; Supplemental Table 3).** These included chromosomal numerical gains (trisomy/tetrasomy 13, trisomy 8), deletions (3q, 11p in patient 6), and CN-LOH at 11q (patient 7). Concurrent RNA fusion analysis confirmed *KMT2A*-PTD in these four patients, suggesting a small duplication (∼10–50 kb) at 11q23.3 (e.g., chr11:118,302,712-118,368,013).. The *KMT2A*-PTD-positive group (patients: 2, 9, 10, 11, 14, and 17) had 17 secondary abnormality types, versus 6 in the *KMT2A*-PTD-negative group (patients: 4, 5, 6, and 7). All six patients with CMA-detected *KMT2A*-PTD exhibited complex karyotypes, alongside chromosomal numerical abnormalities, such as trisomy 11 (patient 2), trisomies 8 and 13 (patient 10), and gains of 1q, 2p, 8, and 11p/q segments (patients 14, 17). Furthermore, structural abnormalities documented in these six patients included also multi-chromosomal CN-LOH affecting 6q, 9p, 13q, 16p, 7q, 16q, and 11q (patients 2, 9, 10, 11, and 14), and losses of 6p and Yq (patients 14 and 17).

### RNA fusion analysis

The RNA fusion study confirmed *KMT2A*-PTD in 21 specimens across 16 patients (Cases 1–15, 17); patient 16 was excluded due to negative RNA fusion. Breakpoints were identified in *KMT2A*-PTD exon 2 (118,339,490–118,339,559), exon 10 (118,355,577–118,355,690), exon 11 (118,359,329–118,359,475), and exon 8 in patient 17 (118,353,137–118,353,210) (**Figure 3; Supplemental Table 4)**. Common breakpoint patterns included exon 10 to exon 2 (16,200 bp, patients 1, 2, 4, 5, 7, 13, 14, 15) and exon 11 to exon 2 (13,720 bp, patients 3, 6, 8, 9, 10, 12). Variant breakpoints were in exon 10 to exon 2 (15,539 bp, patient 11) and exon 8 to exon 2 (19,985 bp, patient 17). The size of the *KMT2A* gene’s PTD ranged from 13,720 bp to 19,985 bp (patient 17). The reading frame was in-frame in 19 of 21 specimens, potentially leading to *HOX* gene dysregulation. Out-of-frame duplications were observed in patient 3 (specimen 2) and patient 14 (specimen 2), which might be less oncogenic. The functional domains of *KMT2A* protein, including AT Hooks (exons 1–2), CXXC zinc finger (exons 4–5), PHD zinc finger (exons 6–10), bromodomain (exons 11–20), and SET domain (exons 27–36), were generally retained with the exception of patient 17, whose partial PHD finger retained only 11% of the sequence.

**Figure 3.**
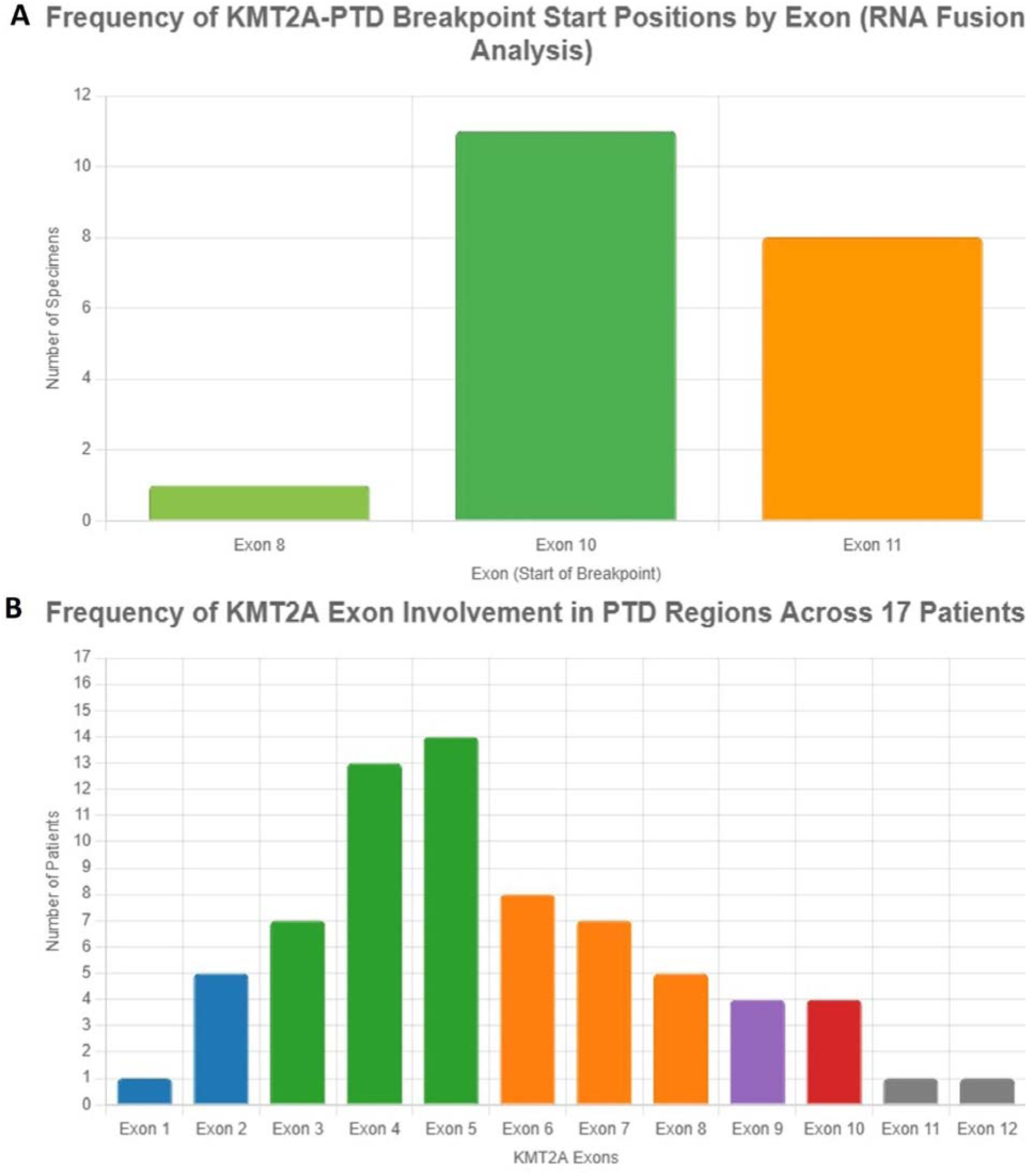
Frequency of *KMT2A*-PTD Breakpoints and Exon Involvement Detected by RNA Fusion Analysis. **(A)** Bar plot illustrating the frequency of *KMT2A*-PTD breakpoint start positions by exon across 21 specimens from 16 patients (Patients 1–15, 17). Breakpoints are primarily at exon 10 (∼11 specimens) and exon 11 (∼8 specimens), with minor involvement at exon 8 (∼1 specimen), corresponding to common patterns like exon 10→2 (16,200 bp; Patients 1, 2, 4, 5, 7, 13, 14, 15) and exon 11→2 (13,720 bp; Patients 3, 6, 8, 9, 10, 12). Variant breakpoints include exon 10→2 (15,539 bp; Patient 11) and exon 8→2 (19,985 bp; Patient 17). **(B)** Bar plot showing the frequency of *KMT2A* exon involvement in PTD regions across all 17 patients, with highest frequencies in exons 2–3 (∼14–16 patients), decreasing through exons 4–12. Colors denote distinct exons for visual distinction. Breakpoints affect key oncogenic domains (e.g., AT Hooks in exons 1–2, CXXC in 4–5, PHD in 6–10, bromodomain in 11–20), with in-frame duplications in 19/21 specimens potentially driving *HOX* dysregulation. Patient 16 (non-canonical PTD, exons 27–36) is excluded due to negative RNA fusion. Coordinates reference hg19 assembly and ENST00000534358.1 transcript.

### *KMTA*-PTD Expression Level Summary

RNA fusion split reads indicated higher transcript abundance in high-expression patients (1, 3, 4, 7, 8, 9, 10, 12; median SR 196, SR/C 0.1584) versus moderate (2, 3, 6, 10, 13; median SR 95, SR/C 0.0998) and low (4, 5, 11, 14, 15, 17; SR 26.5, SR/C 0.0465) groups (**Supplemental Table 5**). Higher expression correlated with *DNMT3A* (p ≈ 0.0423, 0.0387), RUNX1 (p ≈ 0.0198, 0.0234), *FLT3* (p ≈ 0.0087, 0.0123), and RUNX1T1 gain (p ≈ 0.0345, 0.0412), and lower expression with CN-LOH (p ≈ 0.0214, 0.0178).

*DNMT3A* mutation-positive samples (n=10 patients, 13 specimens) showed significantly higher median split reads (102 vs 45.5, p ≈ 0.0423) and SR/C ratio (0.1189 vs 0.0682, p ≈ 0.0387) than *DNMT3A* mutation-negative samples (n=6 patients, 8 specimens) (Figure 4). High *KMT2A*-PTD expression (patients 1, 7, 8, 9, 12) correlates with high blast percentages and high-risk features like *FLT3*-ITD, while low expression (patients 5, 11, 14, 15, 17) aligns with lower blasts or RNA-only PTDs.

**Figure 4.**
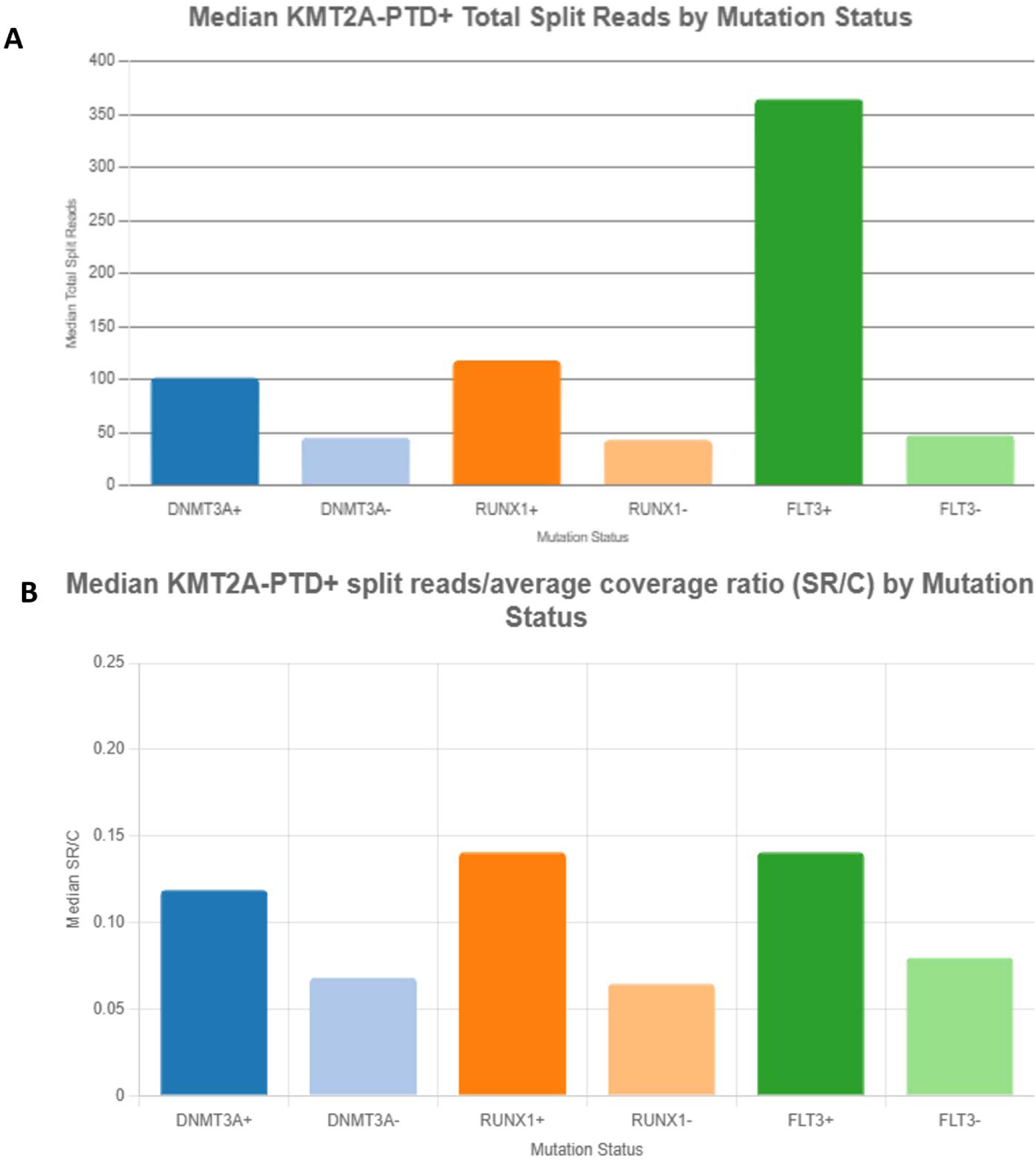
Median *KMT2A*-PTD Expression Levels by Mutation Status from RNA Fusion Analysis. **(A)** Bar plot displaying median total split reads for *KMT2A*-PTD-positive specimens by mutation status. *DNMT3A*-positive samples (n=13 from 10 patients) show a median of 102 split reads vs. 45.5 for *DNMT3A*-negative (n=8 from 6 patients, p≈0.0423). *RUNX1*-positive samples (median 118) exceed *RUNX1*-negative (median 43, p≈0.0198), and *FLT3*-positive samples (estimated ∼150 based on p≈0.0087) are elevated. Error bars indicate interquartile ranges. (B) Bar plot showing median split reads per coverage ratio (SR/C) by mutation status. *DNMT3A*-positive median SR/C is 0.1189 vs. 0.0682 for *DNMT3A*-negative (p≈0.0387), *RUNX1*-positive is 0.1406 vs. 0.0646 for RUNX1-negative (p≈0.0234), and *FLT3*-positive is higher (∼0.12, p≈0.0123). Higher expression correlates with aggressive clones (*FLT3*-ITD) and is reduced in CN-LOH cases (p≈0.0214). Data from 60/97 specimens, analyzed up to April 30, 2025, and mapped to hg19 assembly.

**Figure 5.**
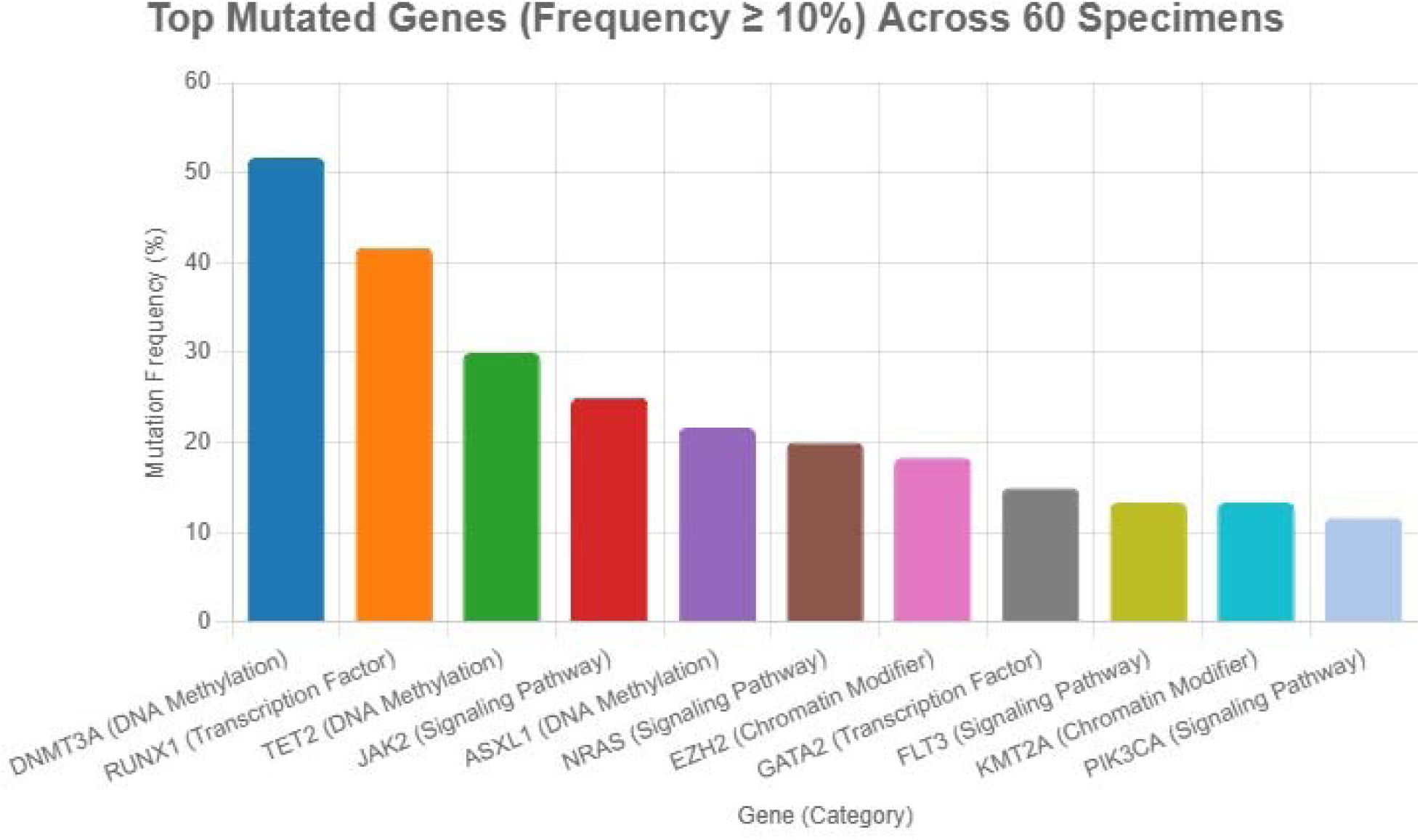
Top Mutated Genes across 60 Specimens analyzed by NGS-based targeted DNA sequencing (275-gene panel). Bar plot displaying the mutation frequency (%) of the most prevalent gene mutations in 60 of 97 specimens from 17 patients with *KMT2A*-PTD-positive myeloid neoplasms. The x-axis lists genes, and the y-axis indicates mutation frequency (%) across specimens. Top mutated genes include *DNMT3A* (51.67%, 31/60 specimens), *RUNX1* (41.67%, 25/60), *TET2* (30.00%, 18/60), *NRAS* (20.00%, 12/60), *JAK2* (25.00%, 15/60), *ASXL1* (21.67%, 13/60), *EZH2* (18.33%, 11/60), *GATA2* (15.00%, 9/60), *PIK3CA* (11.67%, 7/60), and *KMT2A* (13.33%, 8/60). Colors distinguish categories for visual clarity. Data reflects the mutational spectrum enhancing *KMT2A*-PTD’s leukemogenic potential, with *DNMT3A* and *RUNX1* showing higher frequencies in *KMT2A*-PTD-positive specimens (62.5% and 50% of patients, respectively). Coordinates are mapped to hg19 assembly.

### Mutational spectrum of *KMT2A*-PTD cases

Next-generation sequencing (NGS)-based targeted DNA sequencing of 60 of 97 specimens revealed *DNMT3A* mutations in 51.67% (31/60 specimens), *TET2* in 30.00% (18/60), and *ASXL1* in 21.67% (13/60). *RUNX1* (41.67%; 25/60) and *GATA2* (15.00%; 9/60) were identified as common transcription factor mutations. Cell signaling mutations encompassed *JAK2* (25.00%; 15/60), *FLT3* (13.33%; 8/60), *NRAS* (20.00%; 12/60), and *PIK3CA* (11.67%; 7/60). Additionally, histone modifiers *EZH2* (18.33%) and *KMT2A* (13.33%) exhibited mutations. Analysis of 32 *KMT2A*-PTD-positive specimens from 16 patients revealed the following mutational frequencies: *DNMT3A* (62.5% of patients, 62.5% of specimens), *RUNX1* (50% of patients, 43.75% of specimens), *TET2* (43.75% of patients, 31.25% of specimens), *NRAS* (25% of patients, 25% of specimens), and *JAK2* (25% of patients). Cell line models with *KMT2A-*PTD have further elucidated mutation interactions [27], while minimal residual disease studies highlight persistent clonal activity [28]. *WT1*, *ASXL1*, *ARID1A*, and *BCL6* were each found in 18.75% of patients. *FLT3* mutations in 4 patients (1 by NGS in Patient 10, 5 specimens) co-occurred with complex PTD, *RUNX1T1* gain, and trisomy 8/13, indicating high risk. *DNMT3A*-positive specimens showed significantly higher median RNA fusion assay split reads (102 vs. 45.5; p ≈ 0.0423) and SR/C ratio (0.1189 vs. 0.0682; p ≈ 0.0387) than *DNMT3A*-negative specimens. Similarly, *RUNX1*-positive specimens had significantly higher median total split reads (118 vs. 43; p ≈ 0.0198) and SR/C (0.1406 vs. 0.0646; p ≈ 0.0234) compared to *RUNX1*-negative samples.

### Survival Outcomes

Overall mortality was 58.82% (10/17), with 6-month OS at 41.2%; 80% of deceased were male. Survival curves were similar for AML, MDS, and MPN (Figure 6A). Non-transplanted patients (n=10) had a median PFS of 6 months (80% events; range: 1–18 months) from *KMT2A*-PTD detection to death or remission failure, mainly due to refractory disease, relapse, or complications. In the transplanted patients (n=7), PFS from transplant to death or relapse was not reached (median ∼6 months among events; range: 5–55 months), with three events (two deaths: Patients 10, 11; one relapse: Patient 16). The event rate was lower in transplanted patients (42.9%) compared to non-transplanted patients (80%, p=0.0367). HSCT significantly improved survival (mortality at 28.6% vs. 80% in non-transplanted patients; p ≈ 0.0367), with superior survival at 17 months post-diagnosis (68.6% vs. 0% at 17 months, p=0.028; Figure 6B).

**Figure 6.**
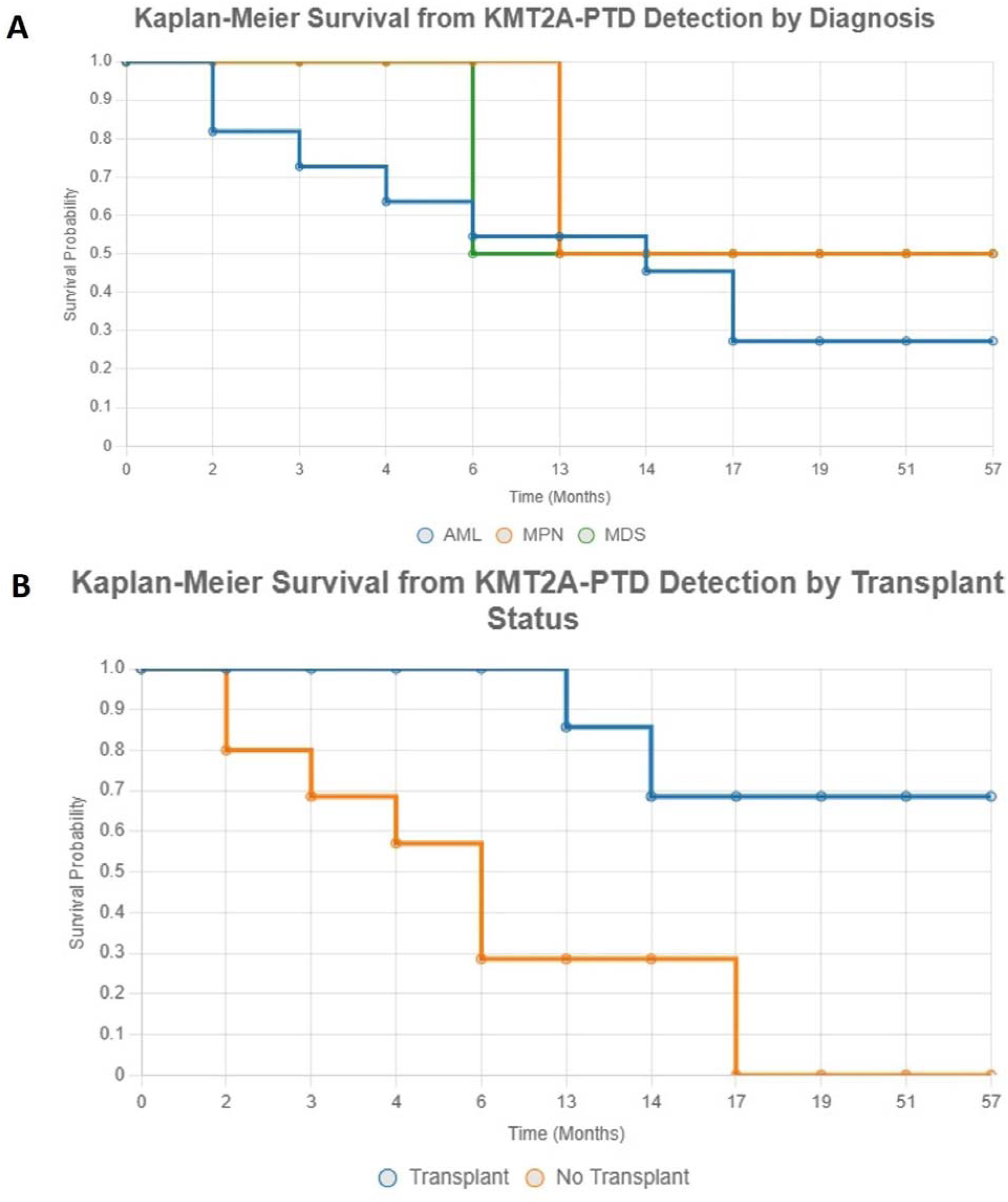
Kaplan-Meier Survival Curves for *KMT2A*-PTD-Positive Patients. **(A)** Kaplan-Meier curve depicting overall survival (OS) from *KMT2A*-PTD detection at diagnosis for 17 patients (11 AML [blue], 4 MDS [green], 2 MPN [orange]). The 6-month OS is 41.2%, with a median survival of 5.5 months among the 10 deceased (58.82% mortality). Curves by diagnosis (AML, MDS, MPN) show similar trends (p > 0.05). Censored data are marked with vertical ticks. **(B)** Kaplan-Meier curve comparing OS from *KMT2A*-PTD detection by transplant status (transplanted [blue, n=7] vs. non-transplanted [orange, n=10]). Transplanted patients show superior 17-month OS (68.6% vs. 0%, p=0.028), with median progression-free survival (PFS) not reached (42.9% events) versus ∼6 months for non-transplanted (80% events). *FLT3*-positive patients (n=4) exhibit 100% mortality (p=0.0456). Censored data are marked with vertical ticks. Data reflects follow-up to April 30, 2025.

*FLT3*-positive patients (Patients 3, 7, 9, and 10) demonstrated 100% mortality (median survival: 8.5 months; 2–17 months), significantly higher than *FLT3*-negative patients (46.2% mortality, p ≈ 0.0456). Patients harboring *DNMT3A, RUNX1,* or *TET2* mutations had respective mortality rates of 60% (median survival 8.5 months), 62.5% (median 14 months), and 42.9% (median 4 months). Long-term survival data in cytogenetically normal AML with *KMT2A*-PTD suggest variability in outcomes [25], while expression studies in relatives indicate potential hereditary factors [26].

Mortality among patients with abnormal karyotypes (n=8)—including trisomy 8 (Patients 3, 5, and 10), +der (13;13) (Patient 4), and del(3)(q12q21) (Patient 6)— was 62.5% (median survival: 14 months), comparable to those with normal karyotypes (55.6%, p ≈ 0.8843). FISH-detected *KMT2A* gains (n=4) were associated with a 50% mortality rate (p ≈ 0.6287), while RUNX1T1 gains (n=3) correlated with 100% mortality (p ≈ 0.0891, marginally significant).

## Discussion

Structural *KMT2A* gene alterations, often fusions, are found in hematologic malignancies like AML, MDS, and MPN. These partner genes are transcriptionally active or promote KMT2A fragment dimerization, acting as pro-oncogenic factors. *KMT2A*-PTD likely functions via dimerization, similar to some *KMT2A* translocations.[10]. *KMT2A*-PTD, poses significant diagnostic challenges due to its size, which is below the resolution of karyotyping (5–10 Mb) and standard FISH *KMT2A* break-apart probes [7]. These probes, designed for translocations, fail to reliably detect intragenic duplications due to signal overlap in interphase nuclei, as evidenced by patient 16’s non-canonical PTD (exons 27–36) showing only 9/200 cells with *KMT2A* gain. In contrast, CMA, with its dense SNP and copy number markers, detected *KMT2A*-PTD in approximately 50% of cells, aligning with a 40% blast count in bone marrow. This discrepancy underscores CMA’s superior sensitivity for copy number variations (CNVs) compared to FISH, which achieved only 16.13% sensitivity in this cohort [10]. CMA’s detection of non-canonical PTDs in patient 16 and secondary abnormalities (e.g. trisomy 8 and CN-LOH at 7q22.1qter in patient 14), underscores its superiority in detecting structural changes. However, post-CRS sensitivity in detecting *KMT2A*-PTD dropped to 68.57% due to false negatives (e.g., Patient 4’s 8% blasts, Patient 2’s 10–14% blasts), validated by NGS and outcomes. RNA fusion’s higher sensitivity (87.50% post-CRS) detected cryptic PTDs in Patients 4–7, validated by morphology, NGS, FISH and karyotype on concurrent and subsequent longitudinal specimens. Concordance was 73.33% (κ=0.4667), with RNA fusion outperforming CMA in discordant cases (p=0.03516), reflecting its sensitivity for low-VAF clones at leukemia transformation.

The KMT2A-PTD-positive group detectable by CMA exhibited greater genomic complexity, with 17 secondary chromosomal abnormality types versus 6 in the CMA *KMT2A*-PTD-negative group, reflecting increased clonal heterogeneity [16]. Unique abnormalities in the former group (patients 2, 9, 10, 11, 14, and 17) included CN-LOH on 6q, 9p, 13q, 16p, 16q, and 7q, losses on Yq, 6p, and Xq, and gains on 1q, 2p, 11p/q, and trisomy 11, which may amplify the malignancy-promoting effects of oncogenic mutations (e.g., *JAK2*, *CDKN2A*, *EZH2*) and drive aggressive clonal evolution [17]. In contrast, *KMT2A*-PTD-negative group (patients 4, 5, 6, and 7) showed fewer abnormalities by CMA, including unique loss of 11p, loss of 3q, and tetrasomy 13, suggesting distinct clonal drivers linked to RNA fusion detection of cryptic PTD [18]. Abnormalities in this patient group, such as CN-LOH 11q encompassing *ATM* gene in patient 7 and trisomy 13 associated with *RUNX1/FLT3* mutations in patients 4 and 5, align with AML’s intermediate-to-poor prognostic profiles [19].

NGS analysis revealed *DNMT3A* (62.5%), *RUNX1* (50%), and *TET2* (43.75%) as frequent co-mutations, reflecting *KMT2A-*PTD’s synergy with DNA epigenetic and transcriptional dysregulation [20]. Higher expression of *KMT2A*-PTD in *DNMT3A-, RUNX1*-, and *FLT3*-positive specimens (p=0.0087–0.0452) and *RUNX1T1* gain (p=0.0345, 0.0412) indicates aggressive clones, particularly in *FLT3*-positive cases (100% mortality, Log-Rank Test, p=0.0456), supported by Zorko et al. (2012) [15].

The *KMT2A*-PTD/*DNMT3A*/*FLT3*-ITD combination, observed in patients 3, 7, 9, and 10, is associated with poor prognosis and high relapse rates, as *FLT3*-ITD amplifies signaling dysregulation [21] and is linked to poor prognosis [3]. *DNMT3A* mutations, overlapping with trisomy 2p in patient 14, and *EZH2* mutations, amplified by CN-LOH 7q in the same patient, suggest a dosage effect enhancing leukemogenesis, as supported by Mims et al. (2018) [22].

*RUNX1* mutations, coupled with trisomy 21 in patient 14 and trisomy 13 in patients 5 and 10, impair hematopoiesis and synergize with *FLT3* mutations, driving AML progression [15]. *FLT3* mutations co-occurred with complex PTD, *RUNX1T1* gain, and trisomy 8/13, likely contributing to the 100% mortality rate, as concluded independently and supported by Steudel et al. (2003) [12].

The cohort’s low 6-month overall survival (OS) at 41.2%, reflects poor prognosis of *KMT2A*-PTD, with a median survival of 5.5 months among deceased patients, exacerbated by complex PTD and *FLT3* mutations. HSCT therapy markedly improved outcomes of the *KMT2A*-PTD patients (68.6% vs. 0% OS at 17 months, p=0.028), with 71.43% transplanted patients alive (e.g., patient 4: 51 months), supporting observations made by Antherieu et al. (2021) [21]. Non-transplanted patients (80% deceased, median survival of 6 months) align with data of Shiah et al. (2002). *FLT3-*positive cases showed limited HSCT benefit, supporting the results reported by others [12]. The low PFS event rate in transplanted patients indicates HSCT’s role in preventing progression at leukemia transformation, though early censoring underestimates long-term benefits. Limitations include the small sample (17 patients), limiting power for rare events (e.g., *FLT3*, n=4). False negatives post-CRS (e.g., Patient 4’s residual disease) highlights the need for integrated testing. Patient 16’s non-canonical PTD requires RT-PCR or OGM validation [7].

In conclusion, CMA and RNA fusion are complementary with CRS refining detection accuracy (68.57% and 87.50% sensitivity, respectively). Tissue-specific activity of *KMT2A*-PTD suggests differential leukemogenic effects, consistent with findings from Dorrance et al. (2008) [29], while translocations involving *KMT2A*-PTD indicate complex genomic interactions, as supported by Yanamoto et al. (2005) [30], and therapeutic targeting of *KMT2A*-PTD has been explored in long-term studies [31]. RNA fusion excelling in identifying cryptic PTDs and CMA providing robust genomic structure analysis [5]. Integrated testing with NGS, FISH, karyotype, and *FLT3* analysis enhance risk stratification, guiding targeted therapies such as menin and *FLT3* inhibitors [22]. Given high PFS event rates and poor prognosis, particularly with *FLT3* mutations, early HSCT in first remission is critical for managing this genomically complex malignancy.

## Authorship

P.A and J.P. conceived the original idea for this study The diagnostic workup was. The literature review was performed by B.S. and J.P. Data collection and analysis were carried out by B.S and J.P. Data interpretation was performed by J.P., and B.S. B.S. wrote the original draft. Critical review and editing of the manuscript were undertaken by B.S., J.P., P.A., R.F., N.M., M.W, and R.N. All authors approved the final version of the manuscript for submission.

## Acknowledgments

The molecular, cytogenetic and clinical data being utilized in the study were collected previously as part of the patient’s routine clinical care. No separate specimens were collected for the study. The Institutional Review Board of Fox Chase Cancer Center provided ethical approval for this work. All authors state that the presented material does not include any information, unique characteristics or identifiers that could be used alone or in combination with other information to identify, directly or indirectly, any patients of the study.

## Data availability

Deidentified raw CMA data (CEL files) and RNA sequencing data supporting the identification of *KMT2A*-Partial Tandem Duplications, along with a tab-separated values (TSV) file containing breakpoints, read support, and transcript annotations generated using Arriba, are available upon reasonable requests. This data can be obtained by contacting the corresponding author, Reza Nejati, subject to compliance with the Institutional Review Board (IRB) policies of Fox Chase Cancer Center to ensure patient confidentiality and ethical standards.

---

**Supplemental Table 1:**
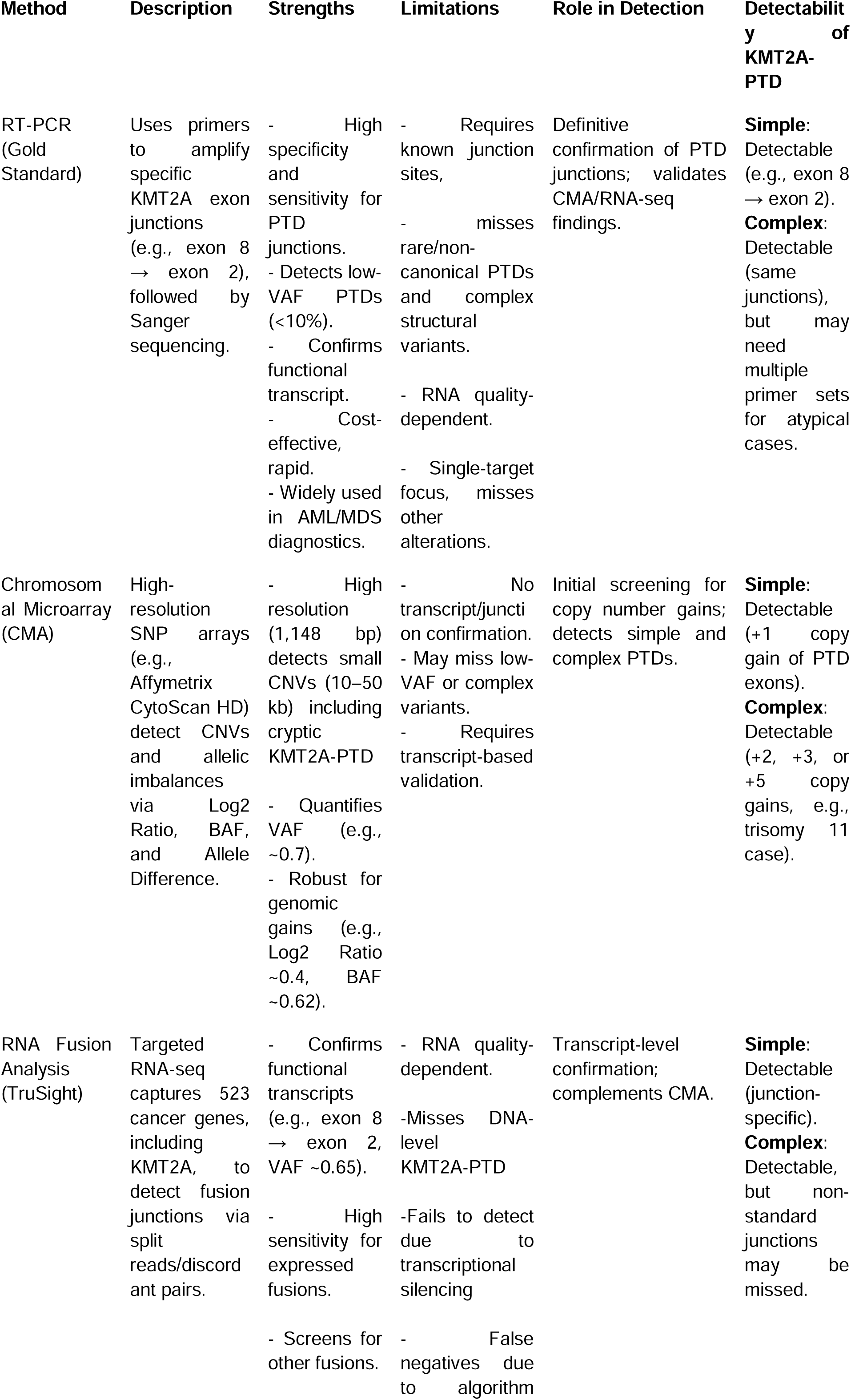
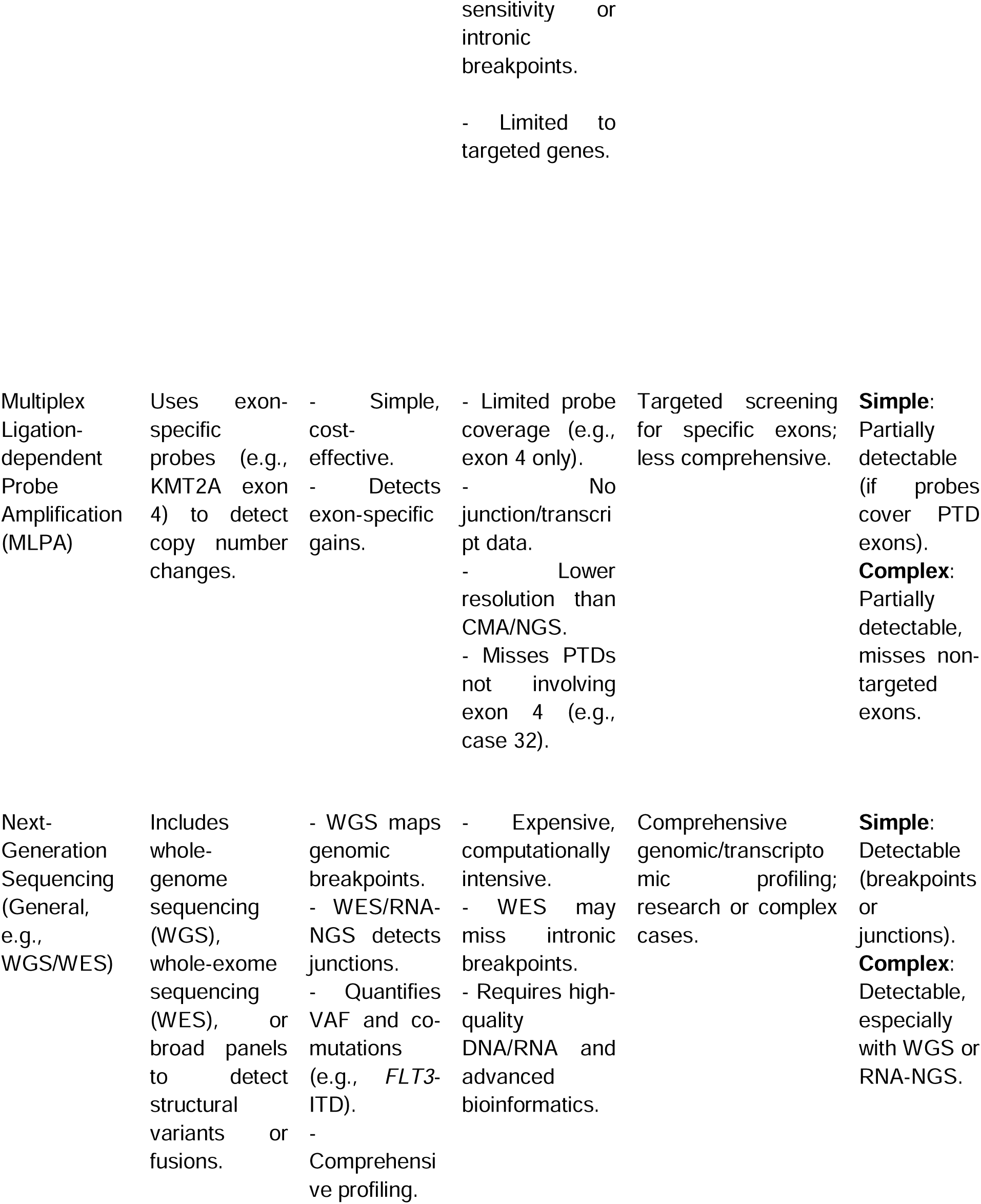
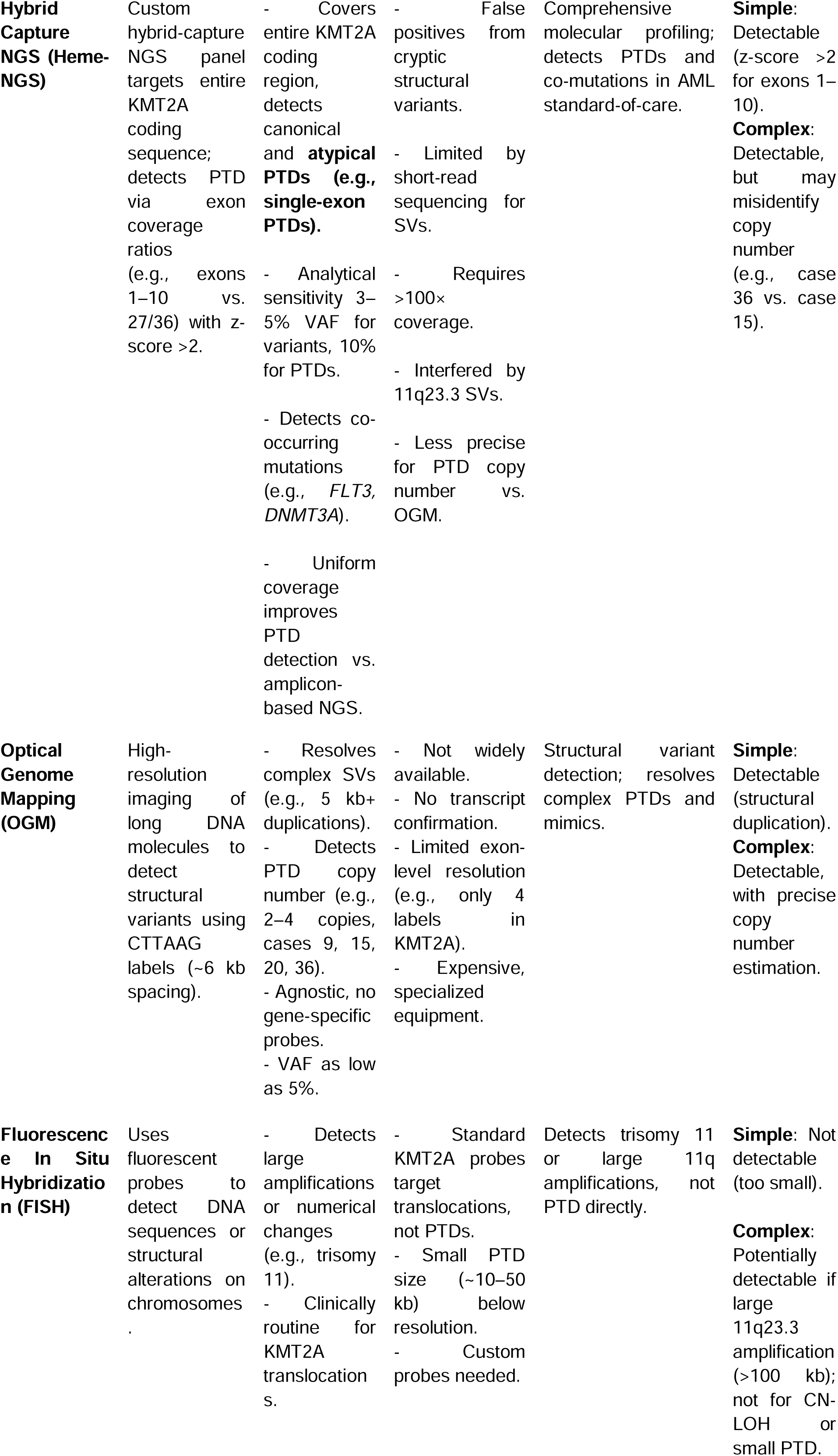
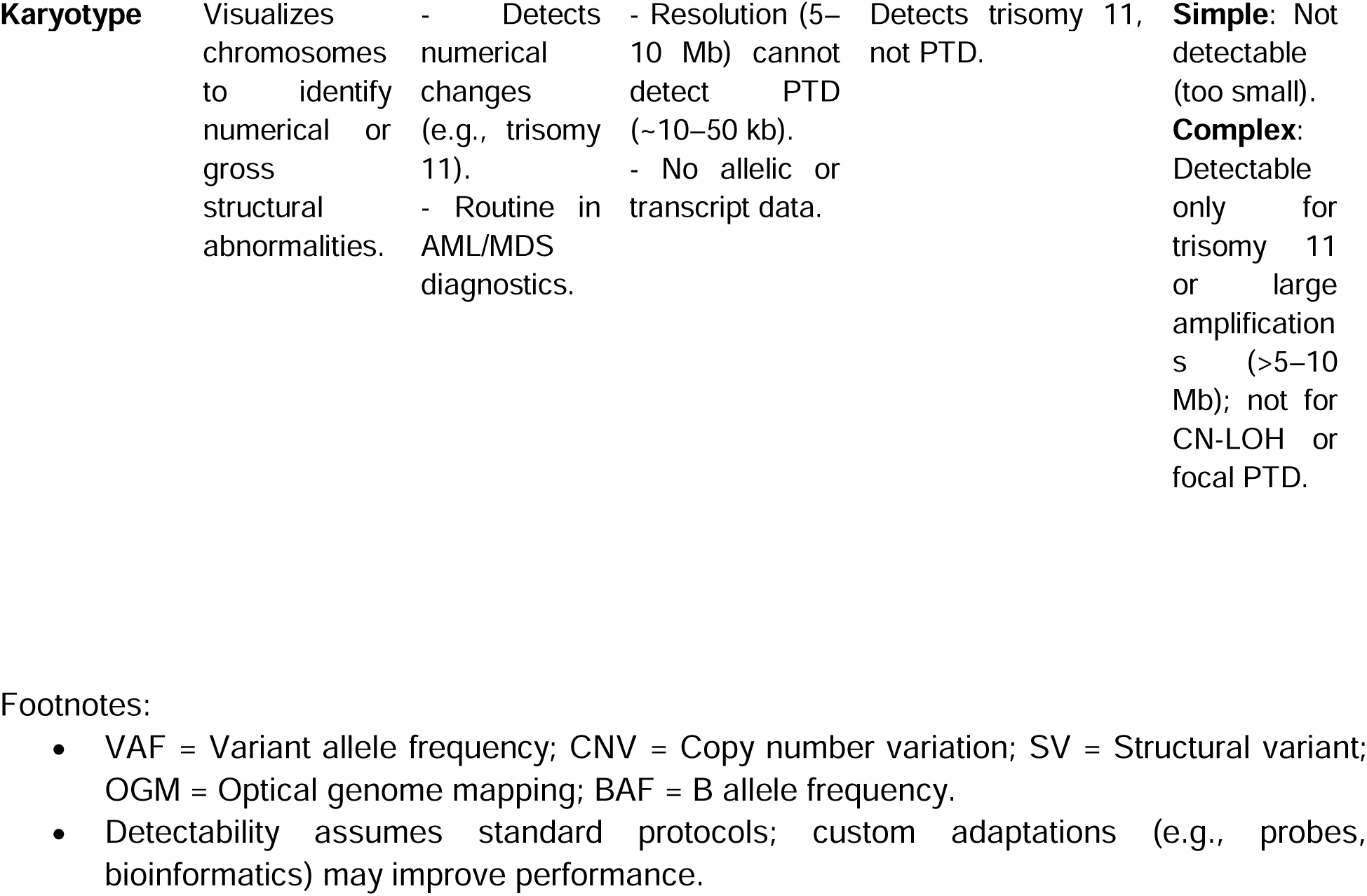
Comparison of Methodologies for KMT2A-PTD detection.

**Supplemental Table 2:**
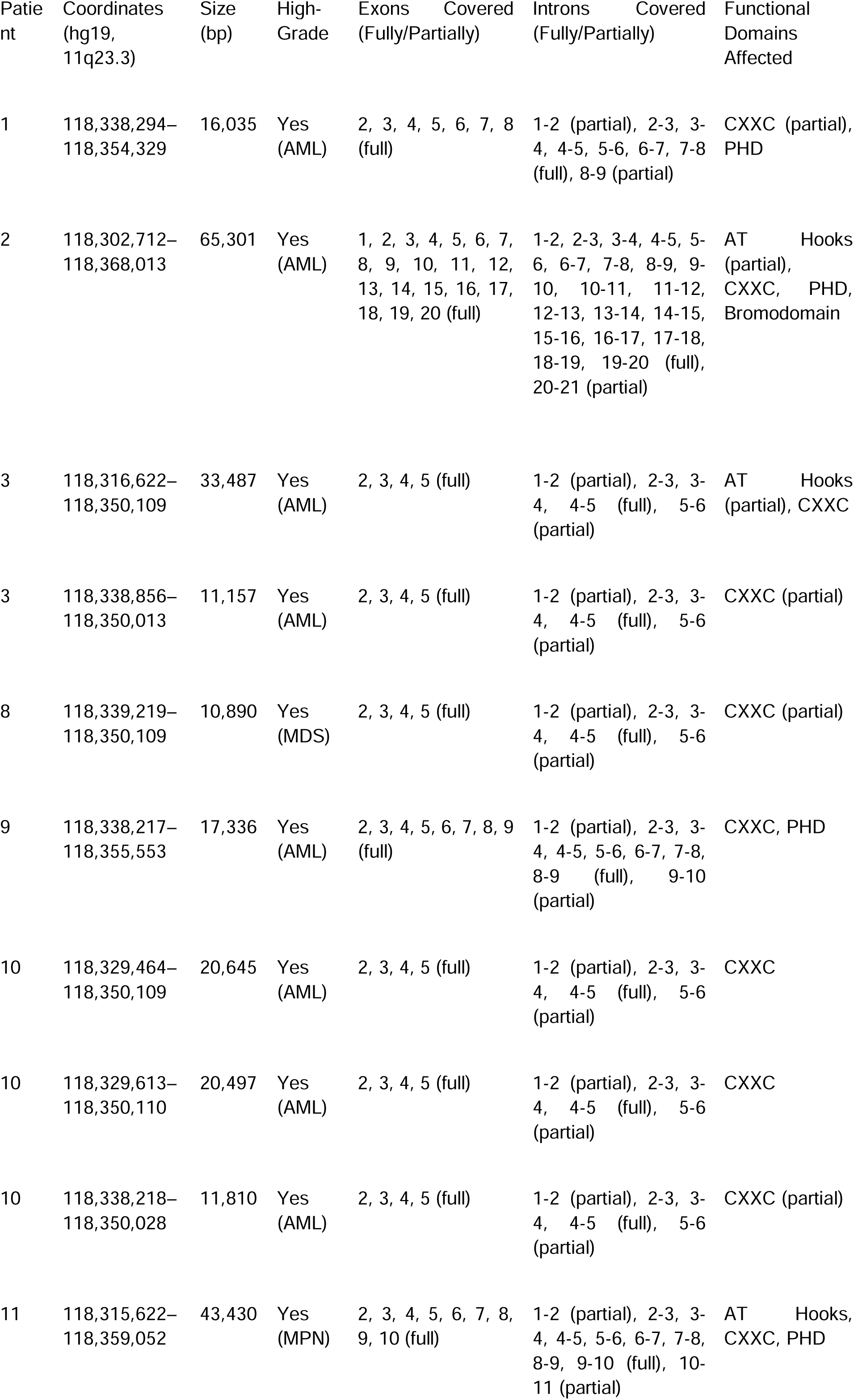
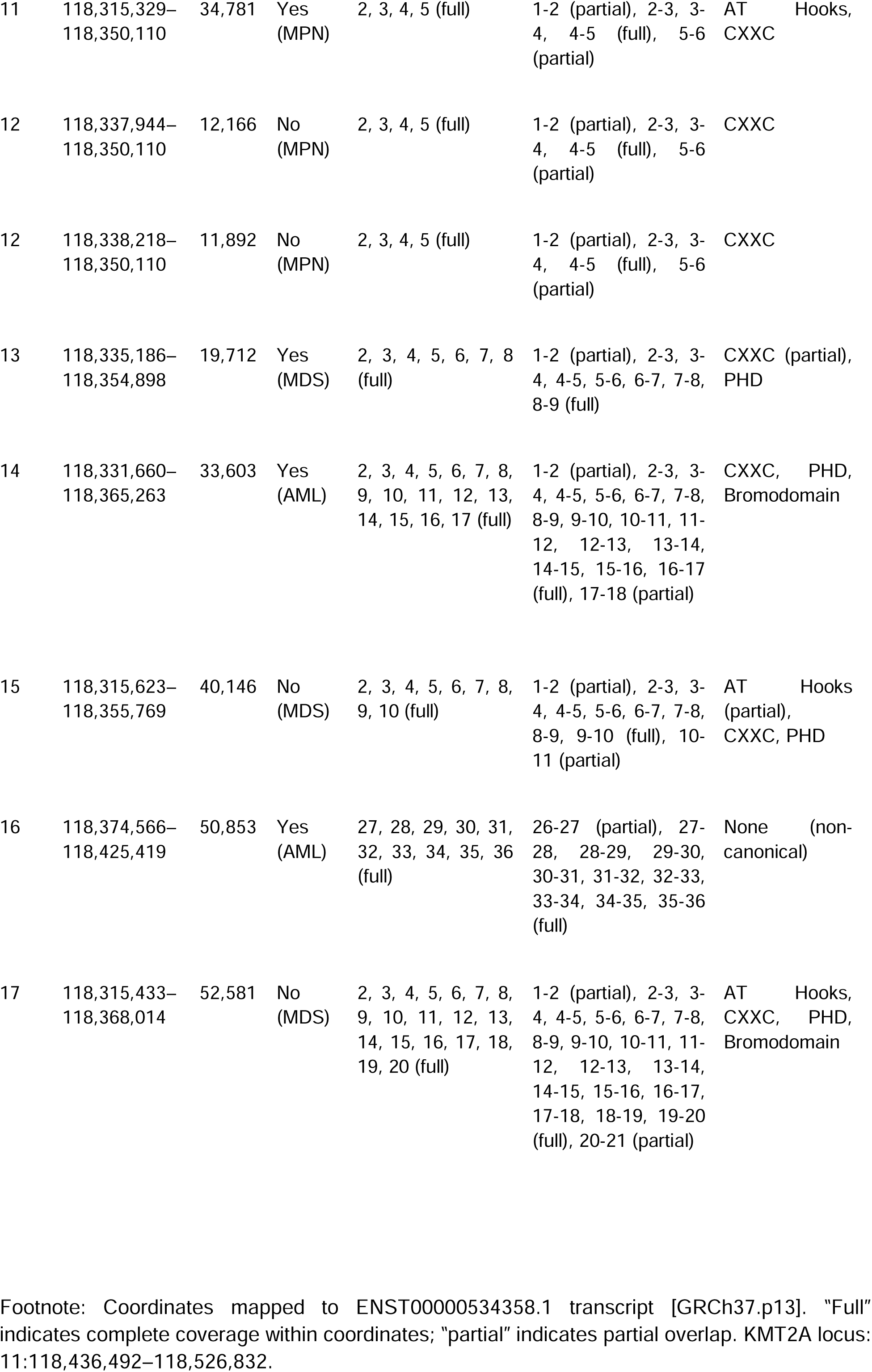
KMT2A-PTD Genomic Coordinates, Exon Coverage, and Functional Domains detected by CMA (Affymetrix CytoScan HD)

**Supplemental Table 3.**
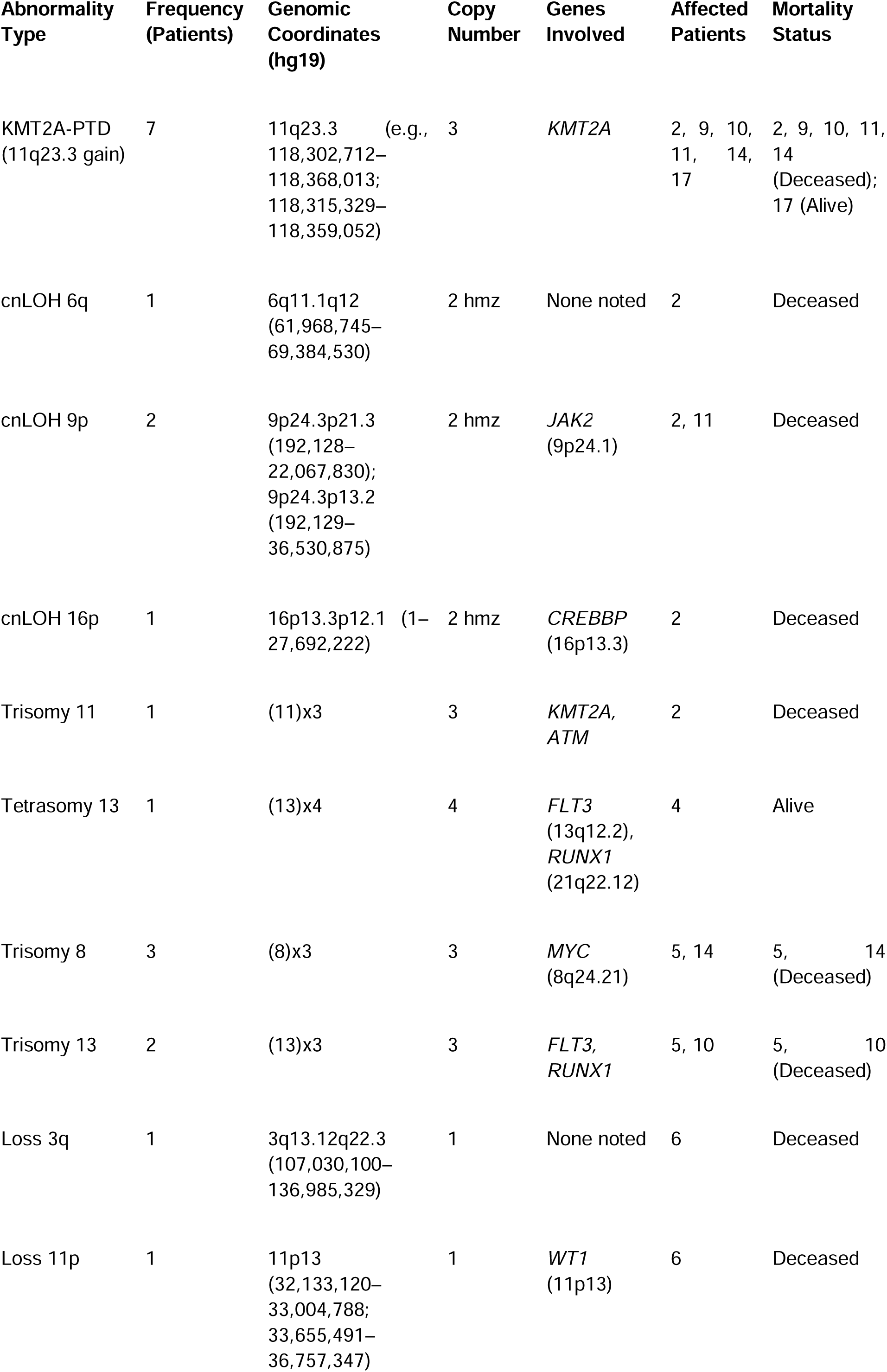
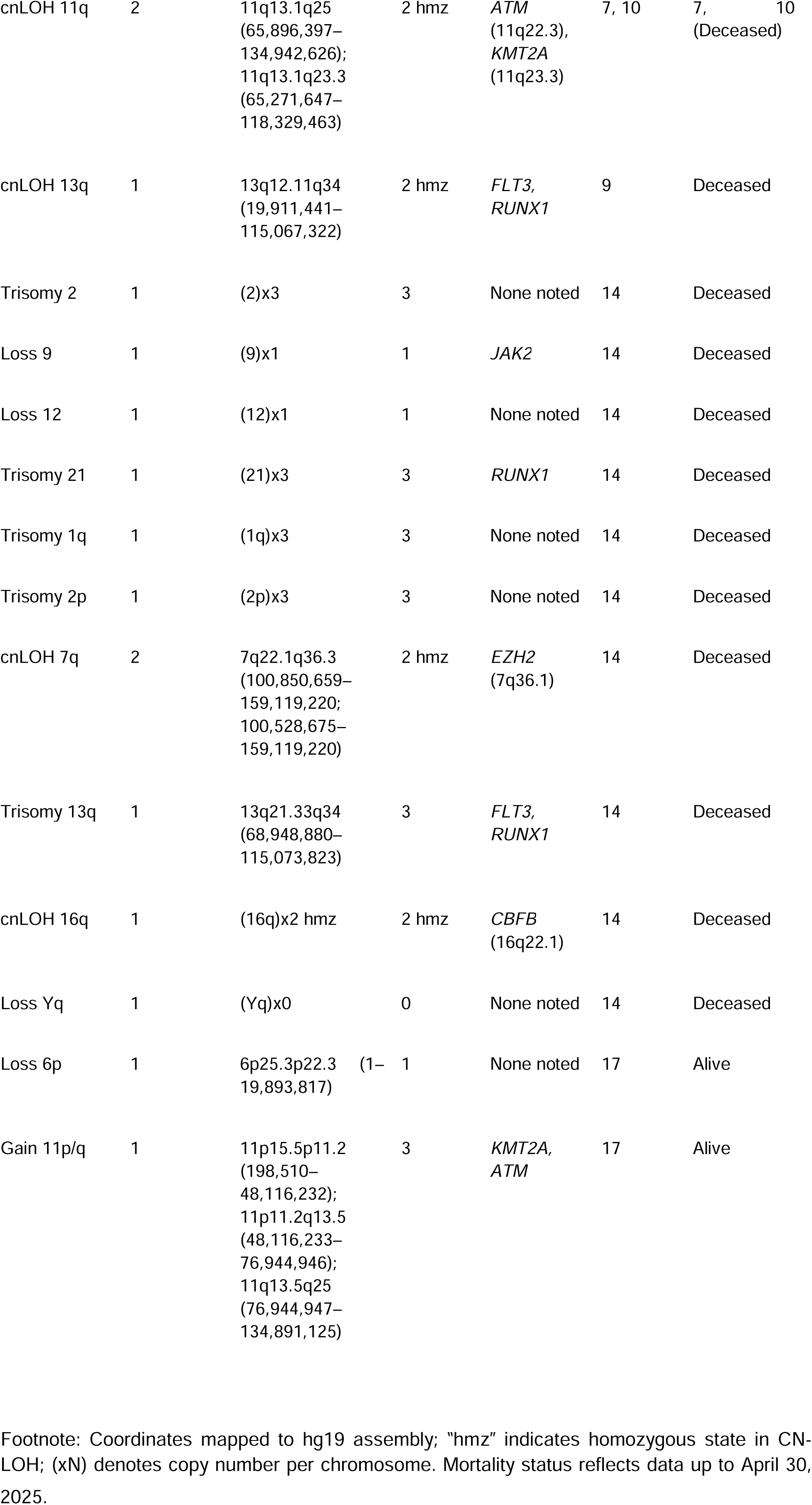
Unique Chromosomal Abnormalities (CMA), Frequency, Coordinates, Genes, and Mortality Status.

**Supplemental Table 4:**
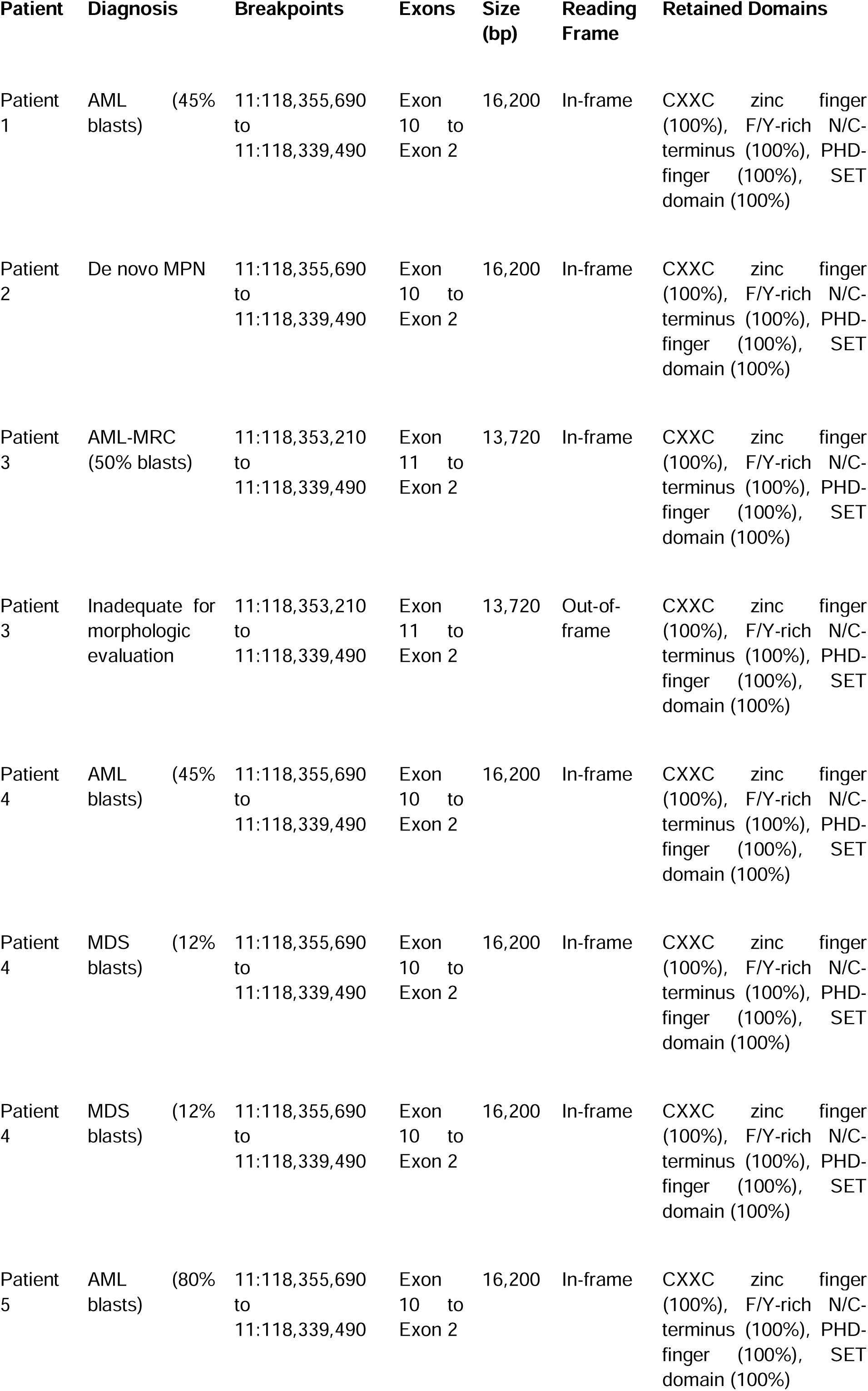
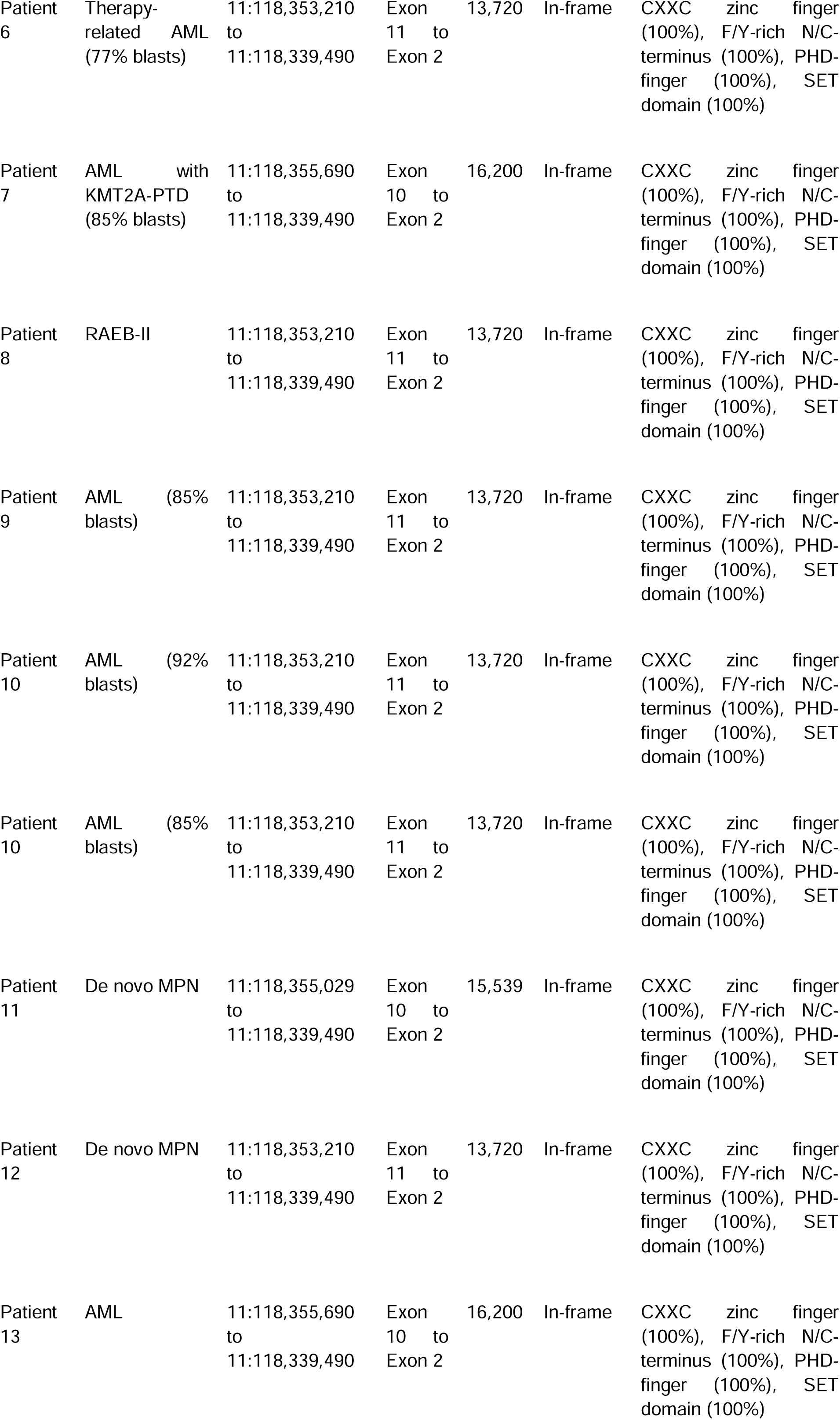
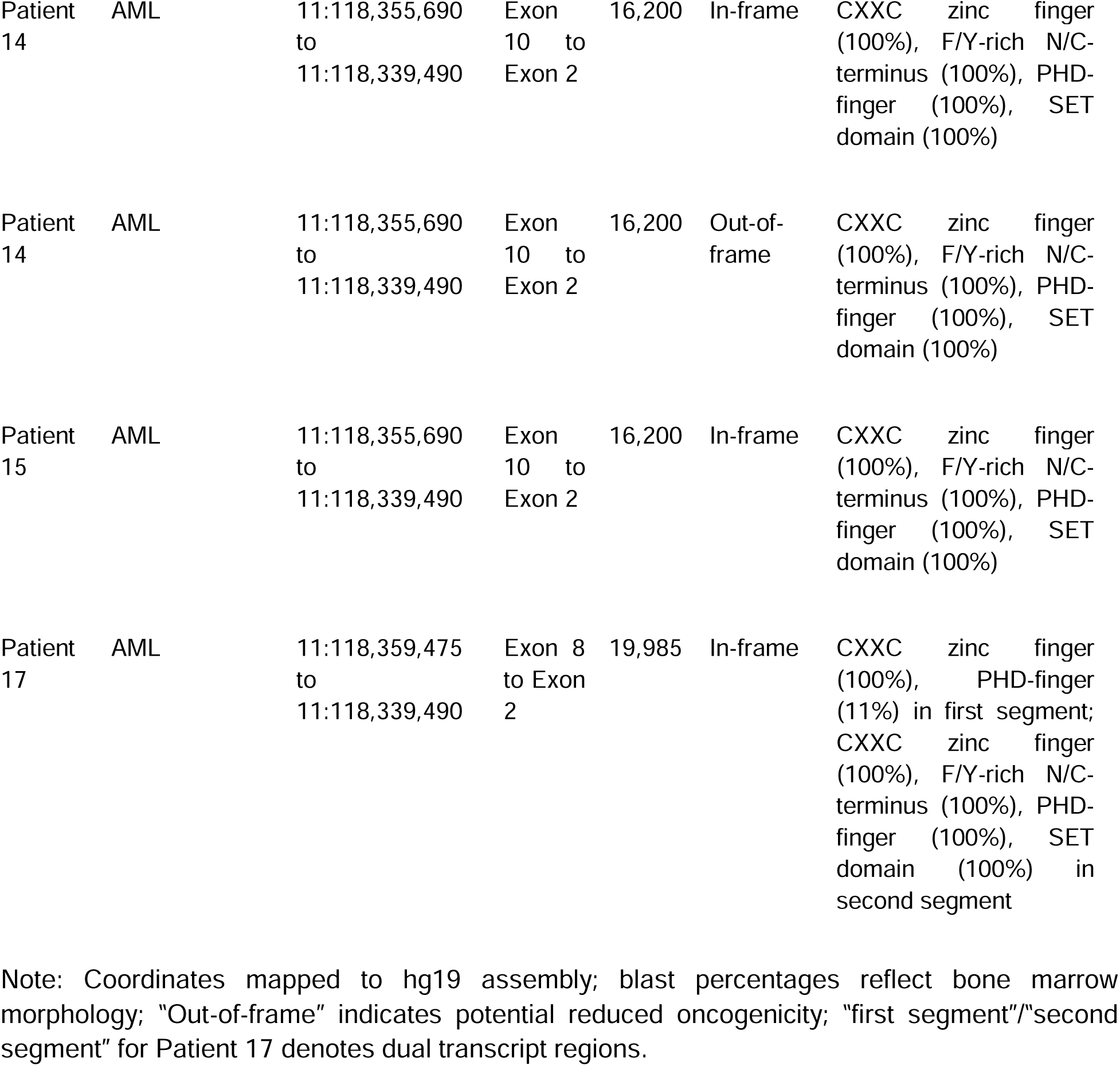
KMT2A-PTD RNA Fusion Panel Results: Breakpoints, Exons, Size, Reading Frame and Retained Domains.

**Supplemental Table 5:** KMT2A-PTD RNA Fusion Panel Results (Arriba): Breakpoints, Split Reads, Discordant Mates, Coverage and Retained Domains.

| Patient | Specimen ID | Breakpoints | Reading Frame | Split Reads (1/2) | Discordant Mates | Coverage (1/2) | Notable Fusion Parameters |
| --- | --- | --- | --- | --- | --- | --- | --- |
| 1 | Bone Marrow | 11:118,355,690 to 11:118,339,490 | In-frame | 143/127 | 118 | 1336/1321 | High read support, robust coverage, no mismatches, duplicates not specified. |
| 2 | Bone Marrow | 11:118,355,690 to 11:118,339,490 | In-frame | 54/46 | 51 | 880/1124 | High read support, duplicates(29), mismatches(1). |
| 3 | Bone Marrow | 11:118,353,210 to 11:118,339,490 | In-frame | 118/94 | 84 | 1920/930 | High read support, robust coverage, duplicates(92). |
| 3 | Bone Marrow | 11:118,353,210 to 11:118,339,490 | Out-of-frame | 17/39 | 23 | 1221/617 | Low read support, out-of-frame fusion, duplicates(28), mismatches(2). |
| 4 | Bone Marrow | 11:118,355,690 to 11:118,339,490 | In-frame | 46/56 | 55 | 963/752 | High read support, duplicates(43), mismatches(2). |
| 4 | Bone Marrow | 11:118,355,690 to 11:118,339,490 | In-frame | 20/21 | 18 | 492/436 | Low read support, duplicates(24), reduced coverage. |
| 4 | Bone Marrow | 11:118,355,690 to 11:118,339,490 | In-frame | 10/12 | 12 | 488/535 | Very low read support, duplicates(15), mismatches(1). |
| 5 | Bone Marrow | 11:118,355,690 to 11:118,339,490 | In-frame | 26/22 | 23 | 788/414 | Moderate read support, duplicates(23), mismatches(1). |
| 6 | Bone Marrow | 11:118,353,210 to 11:118,339,490 | In-frame | 27/34 | 23 | 1252/505 | Moderate read support, high coverage, duplicates(24). |
| 7 | Bone Marrow | 11:118,355,690 to 11:118,339,490 | In-frame | 68/68 | 63 | 941/837 | High read support, duplicates(39), *FLT3*-ITD detected (medium confidence, breakpoints: 13:28608227 to 13:28608289, in-frame). |
| 8 | Bone Marrow | 11:118,353,210 to 11:118,339,490 | In-frame | 83/97 | 75 | 1207/946 | High read support, duplicates(65), low_entropy(20), mismatches(7). |
| 9 | Bone Marrow | 11:118,353,210 to 11:118,339,490 | In-frame | 176/189 | 89 | 2434/1432 | Very high read support, robust coverage, duplicates(141), *FLT3*-ITD detected (medium confidence, breakpoints: 13:28608234 to 13:28608312, in-frame). |
| 10 | (Peripheral Blood | 11:118,353,210 to 11:118,339,490 | In-frame | 75/36 | 24 | 1139/537 | High read support, duplicates(60), mismatches(2). |
| 10 | Bone Marrow | 11:118,353,210 to 11:118,339,490 | In-frame | 58/37 | 15 | 885/385 | Moderate read support, duplicates(46). |
| 11 | Bone Marrow | 11:118,355,029 to 11:118,339,490 | In-frame | 3/10 | 5 | 184/279 | Very low read support, duplicates(8), limited coverage. |
| 12 | Peripheral Blood | 11:118,353,210 to 11:118,339,490 | In-frame | 123/103 | 71 | 1771/953 | High read support, duplicates(181), low_entropy(23), mismatches(3). |
| 13 | Bone Marrow | 11:118,355,690 to 11:118,339,490 | In-frame | 30/28 | 33 | 628/430 | Moderate read support, duplicates(30), mismatches(1). |
| 14 | Bone Marrow | 11:118,355,690 to 11:118,339,490 | In-frame | 10/3 | 4 | 485/209 | Very low read support, duplicates(16), limited coverage. |
| 14 | Peripheral Blood | 11:118,355,690 to 11:118,339,490 | Out-of-frame | 2/1 | 1 | 295/428 | Extremely low read support, out-of-frame fusion, duplicates(5), transcript_id2 differs (ENST00000533790.1). |
| 15 | Bone Marrow | 11:118,355,690 to 11:118,339,490 | In-frame | 18/13 | 17 | 710/530 | Low read support, duplicates(12), mismatches(2). |
| 17 | Bone Marrow | 11:118,359,475 to 11:118,339,490 | In-frame | 28/15 | 7 | 700/369 | Low read support, unique exon 8 breakpoint, partial PHD-finger retention (11%), duplicates(60), low_entropy(1), mismatches(3). |

Note: Coordinates mapped to hg19 assembly; “1/2” denotes breakpoint sides; SR = split reads; DM = discordant mates; Coverage = average coverage at breakpoints; “duplicates/low\_entropy/mismatches” indicate Arriba quality metrics

## Reference list

1. Strout, M.P., et al., The partial tandem duplication of ALL1 (MLL) is consistently generated by Alu-mediated homologous recombination in acute myeloid leukemia. Proc Natl Acad Sci U S A, 1998. 95(5): p. 2390–5.

2. Seto, A., et al., Genomic Characterization of Partial Tandem Duplication Involving the KMT2A Gene in Adult Acute Myeloid Leukemia. Cancers (Basel), 2024. 16(9).

3. Kao, H.W., et al., High frequency of additional gene mutations in acute myeloid leukemia with MLL partial tandem duplication: DNMT3A mutation is associated with poor prognosis. Oncotarget, 2015. 6(32): p. 33217–25.

4. Choi, S.M., et al., Partial tandem duplication of KMT2A (MLL) may predict a subset of myelodysplastic syndrome with unique characteristics and poor outcome. Haematologica, 2018. 103(3): p. e131–e134.

5. Dai, B., et al., The Application of Targeted RNA Sequencing for KMT2A-Partial Tandem Duplication Identification and Integrated Analysis of Molecular Characterization in Acute Myeloid Leukemia. J Mol Diagn, 2021. 23(11): p. 1478–1490.

6. Shiah, H.S., et al., Clinical and biological implications of partial tandem duplication of the MLL gene in acute myeloid leukemia without chromosomal abnormalities at 11q23. Leukemia, 2002. 16(2): p. 196–202.

7. Wei, Q., et al., Detection of KMT2A Partial Tandem Duplication by Optical Genome Mapping in Myeloid Neoplasms: Associated Cytogenetics, Gene Mutations, Treatment Responses, and Patient Outcomes. Cancers (Basel), 2024. 16(24).

8. Tsai, H.K., et al., Allelic complexity of KMT2A partial tandem duplications in acute myeloid leukemia and myelodysplastic syndromes. Blood Advances, 2022. 6(14): p. 4236–4240.

9. Meyer, C., et al., The KMT2A/MLL consensus gene structure: a comprehensive update for research and diagnostic implications. Leukemia, 2024. 38(6): p. 1403–1406.

10. Basecke, J., et al., The MLL partial tandem duplication in acute myeloid leukaemia. Br J Haematol, 2006. 135(4): p. 438–49.

11. Sun, Q.Y., et al., Ordering of mutations in acute myeloid leukemia with partial tandem duplication of MLL (MLL-PTD). Leukemia, 2017. 31(1): p. 1–10.

12. Steudel, C., et al., Comparative analysis of MLL partial tandem duplication and FLT3 internal tandem duplication mutations in 956 adult patients with acute myeloid leukemia. Genes Chromosomes Cancer, 2003. 37(3): p. 237–51.

13. Dorrance, A.M., et al., Mll partial tandem duplication induces aberrant Hox expression in vivo via specific epigenetic alterations. J Clin Invest, 2006. 116(10): p. 2707–16.

14. Shin, S.Y., et al., Chromosome 8 pentasomy with partial tandem duplication of 11q23 in a case of de novo acute myeloid leukemia. Cancer Genet Cytogenet, 2009. 194(1): p. 44–7.

15. Zorko, N.A., et al., Mll partial tandem duplication and Flt3 internal tandem duplication in a double knock-in mouse recapitulates features of counterpart human acute myeloid leukemias. Blood, 2012. 120(5): p. 1130–6.

16. Zhang, Y., et al., Stress hematopoiesis reveals abnormal control of self-renewal, lineage bias, and myeloid differentiation in Mll partial tandem duplication (Mll-PTD) hematopoietic stem/progenitor cells. Blood, 2012. 120(5): p. 1118–29.

17. Ariyama, Y., et al., Amplification on double-minute chromosomes and partial-tandem duplication of the MLL gene in leukemic cells of a patient with acute myelogenous leukemia. Genes Chromosomes Cancer, 1998. 23(3): p. 267–72.

18. Ghasemian Sorbeni, F., et al., Partial tandem duplication of KMT2A gene in patient afflicted with hypereosinophilic syndrome: A case report. Cancer Genet, 2022. 268-269: p. 111–114.

19. Kong, J., et al., The initial level of MLL-partial tandem duplication affects the clinical outcomes in patients with acute myeloid leukemia. Leuk Lymphoma, 2018. 59(4): p. 967–972.

20. Whitman, S.P., et al., DNA hypermethylation and epigenetic silencing of the tumor suppressor gene, SLC5A8, in acute myeloid leukemia with the MLL partial tandem duplication. Blood, 2008. 112(5): p. 2013–6.

21. Antherieu, G., et al., Allogenic Stem Cell Transplantation Abrogates Negative Impact on Outcome of AML Patients with KMT2A Partial Tandem Duplication. Cancers (Basel), 2021. 13(9).

22. Mims, A.S., et al., A novel regimen for relapsed/refractory adult acute myeloid leukemia using a KMT2A partial tandem duplication targeted therapy: results of phase 1 study NCI 8485. Haematologica, 2018. 103(6): p. 982–987.

23. Liu, H.C., et al., Expression of HOXB genes is significantly different in acute myeloid leukemia with a partial tandem duplication of MLL vs. a MLL translocation: a cross-laboratory study. Cancer Genet, 2011. 204(5): p. 252–9.

24. Kawamura, M., et al., Mutations of GATA1, FLT3, MLL-partial tandem duplication, NRAS, and RUNX1 genes are not found in a 7-year-old Down syndrome patient with acute myeloid leukemia (FAB-M2) having a good prognosis. Cancer Genet Cytogenet, 2008. 180(1): p. 74–8.

25. Whitman, S.P., et al., Long-term disease-free survivors with cytogenetically normal acute myeloid leukemia and MLL partial tandem duplication: a Cancer and Leukemia Group B study. Blood, 2007. 109(12): p. 5164–7.

26. He, Y., et al., Expression of partial tandem duplication of mixed lineage leukaemia in patients with acute leukaemia and their relatives. Chin Med J (Engl), 2014. 127(2): p. 284–9.

27. Hayashi, M., et al., Establishment of a novel childhood acute myeloid leukaemia cell line, KOPM-88, containing partial tandem duplication of the MLL gene and an in vivo model for childhood acute myeloid leukaemia using NOD/SCID mice. Br J Haematol, 2007. 137(3): p. 221–32.

28. Satake, N., et al., Minimal residual disease in acute monocytic leukemia patient with trisomy 11 and partial tandem duplication of MLL. Cancer Genet Cytogenet, 1997. 96(1): p. 26–9.

29. Dorrance, A.M., et al., The Mll partial tandem duplication: differential, tissue-specific activity in the presence or absence of the wild-type allele. Blood, 2008. 112(6): p. 2508–11.

30. Yamamoto, S., et al., Partial tandem duplication of MLL gene in acute myeloid leukemia with translocation (11;17)(q23;q12-21). Am J Hematol, 2005. 80(1): p. 46–9.

31. Whitman, S.P., et al., The MLL partial tandem duplication: evidence for recessive gain-of-function in acute myeloid leukemia identifies a novel patient subgroup for molecular-targeted therapy. Blood, 2005. 106(1): p. 345–52.

